# Using machine learning and centrifugal microfluidics at the point-of-need to predict clinical deterioration of patients with suspected sepsis within the first 24 h

**DOI:** 10.1101/2024.10.08.24314844

**Authors:** Lidija Malic, Peter G.Y. Zhang, Pamela Plant, Liviu Clime, Christina Nassif, Dillon Da Fonte, Evan E. Haney, Byeong-Ui Moon, Victor Sit, Daniel Brassard, Maxence Mounier, Eryn Churcher, James T. Tsoporis, Reza Falsafi, Manjeet Bains, Andrew Baker, Uriel Trahtemberg, Ljuboje Lukic, John C. Marshall, Matthias Geissler, Robert E.W. Hancock, Teodor Veres, Claudia C. dos Santos

## Abstract

Sepsis is the body’s dysfunctional response to infection associated with organ failure. Delays in diagnosis have a substantial impact on survival. Herein, samples from 586 in-house patients were used in conjunction with machine learning and cross-validation to narrow a gene expression signature of immune cell reprogramming to predict clinical deterioration in patients with suspected sepsis within the first 24 hours (h) of clinical presentation using just six genes (Sepset). The accuracy of the test (∼90% in early intensive care unit (ICU) and 70% in emergency room patients) was validated in 3,178 patients from existing independent cohorts. A real-time reverse transcriptase polymerase chain reaction (RT-PCR)-based test was shown to have a 98% sensitivity in >230 patients to predict worsening of the sequential organ failure scores or admission to the ICU within the first 24 h following Sepset detection. A stand-alone centrifugal microfluidic instrument that integrates the entire automated workflow for detection of the Sepset classifier in whole blood using digital droplet PCR was developed and tested. This PREcision meDIcine for CriTical care (PREDICT) system had a high sensitivity of 92%, specificity of 89%, and an overall accuracy of 88% in identifying the risk of imminent clinical deterioration in patients with suspected sepsis.

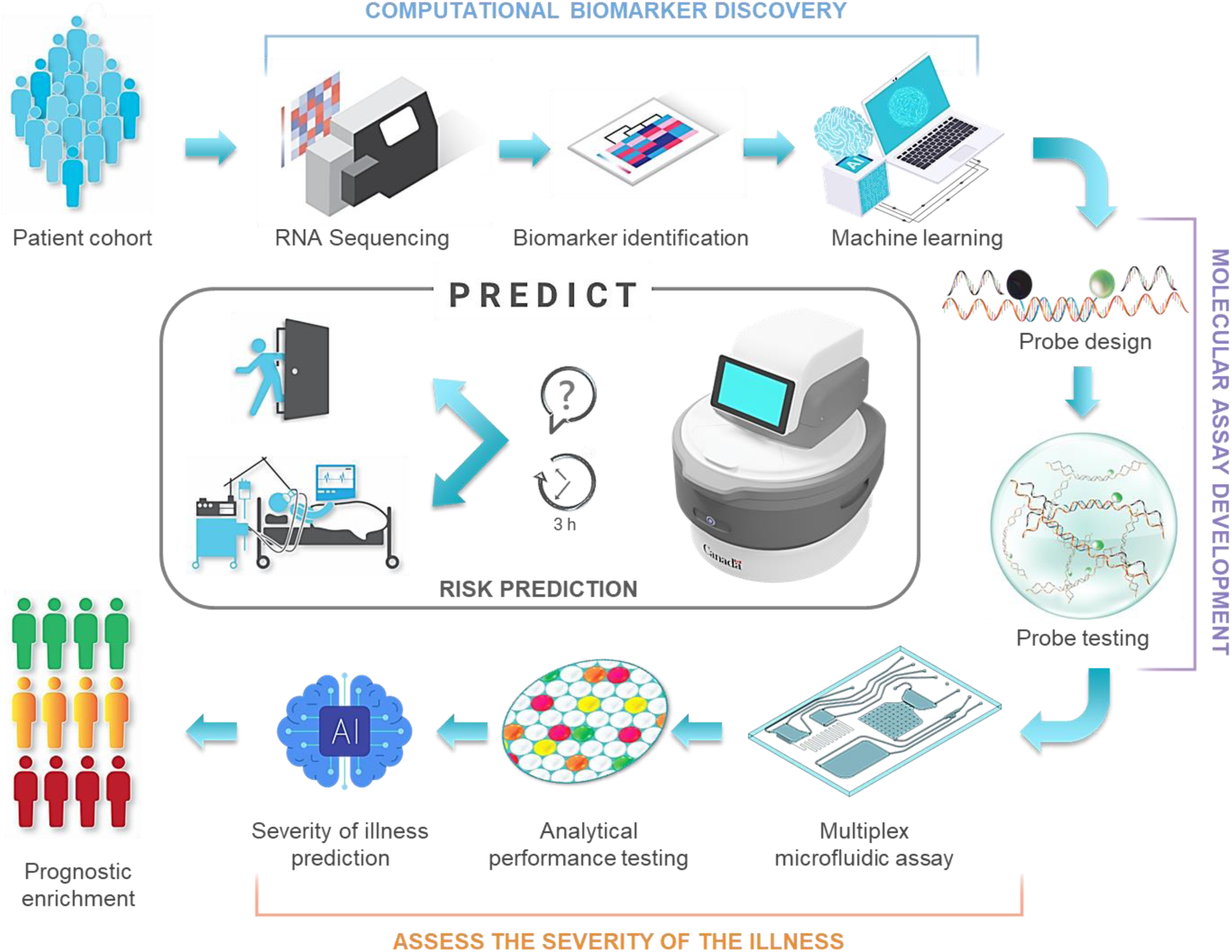
Description of Graphic Abstract. Feature reduction and development of a gene classifier that predicts deterioration-risk-groups in patients starts with in-house RNA sequencing data from patient collected from a heterogenous cohort of patients with suspected sepsis (top left) to reduce our original published gene signature down to 6-genes (Sepset), for which expression could be related to 2 housekeeping genes. Feature selection was performed using machine learning (ML) and AI and the classifier validated in samples from published transcriptomic studies. Molecular assay is then developed by designing and testing primer/probe sequences specific to the target genes using digital droplet PCR. In parallel, sample-to-answer microfluidic platform and cartridges are developed (bottom right) and analytical performance of multiplex quantitative assay is tested. Prognostic enrichment is obtained by analyzing the results using ML algorithm to determine the percent likelihood of significant clinical deterioration within the immediate next 24 h. The deployment of PREDICT platform (center) at the point-of-care is anticipated to aid in triage and management of prospective sepsis within the first 3 h of clinical presentation.

## Introduction

Sepsis, a complex syndrome of organ dysfunction caused by a dysregulated host response to infection,^1^ has been declared a global emergency.^2^ Estimates from 2017 are >48.9 million cases and 11.0 million deaths per year,^3^ not including deaths from COVID-sepsis.^4^ Early individualized interventions^5,6^ may significantly reduce mortality and morbidity, and prevent poor long-term outcomes and disability.^5,6,7^ It has been shown that even short delays in appropriate treatment can cause significant increases in mortality from sepsis.^7^ Despite decades of research, however, patients are still triaged and treated on the basis of clinical symptoms.^8^ While these include measures of overall severity, they are largely nonspecific and do not adequately assess dysregulated responses to infection, align patients with appropriate pharmacotherapies, or predict impending deterioration and the need for resuscitative level care. Biomarkers that provide early prognostic and therapeutic enrichment are actively sought to achieve the personalization necessary to improve research and care.^2,9^

Importantly, while some diagnostic tests have moved to a ‘distributed’ model with relevant tests performed at bedside (e.g., glucose monitoring for diabetes), for patients with sepsis, the vast majority of biochemical and molecular tests are implemented in a ‘centralized’ format, found only in well-equipped laboratories that require trained operators and specialized instruments, making them less accessible (or even inaccessible) to some of our most vulnerable and difficult to access populations.^8,9,10,11^ The need to send samples to the lab for analysis significantly delays results (some more than the 6 h recommended by the surviving sepsis guidelines)^12^ and limits the ability to provide timely care to patients.^6,13,14^ This ‘bottleneck’ means that testing standards fail one of the primary objectives of the World Health Organization (WHO) “to promote health, …and serve the vulnerable so everyone, everywhere can attain the highest level of health.”^15^

Lab-on-a-chip (LOC)^16,17,18,19^ technologies have the potential to advance care beyond traditional ‘syndromic’ approaches, outside specialized centers, promising to democratize care for millions of patients.^20,21,22^ In addition to performing assays in a compact, miniaturized format, LOC devices promote portability and point-of-care (POC) testing, enabling minimally trained personnel to perform analytical procedures outside laboratory settings. We have previously shown that extraction and detection of molecular markers from biological samples, such as blood, can be fully automated using a centrifugal-based LOC system.^23^ We have further integrated bioanalytical assays for pathogen detection involving polymerase chain reaction (PCR)^24^ and loop-mediated isothermal amplification in a sample-to-answer format.^25^ Combining our LOC system with innovative RNA-based biomarkers that predict clinical deterioration^26,27,28,29,30^ may significantly advance sepsis care in specialized and non-specialized settings alike.

In parallel, our group discovered and validated a 99 gene signature, present within 2 h of presentation to the emergency department (ED), able to predict clinical deterioration based on the emergence of a cellular reprogramming profile associated, with the inability of cells to respond to pathogens.^31,32^ Our original expression signature was pathogen agnostic, predicting both all-cause and COVID sepsis and different immunological response endotypes. The signature was also predictive of organ dysfunction and severity outcomes.^30,33^ Here, we hypothesized that a reduced gene signature, would retain the ability to discriminate patients with suspected sepsis at high risk of clinical deterioration. The ability of this signature to classify patients defined by Sepsis-3 criteria^1^ was validated in 3,178 patient samples from nine published transcriptomic studies. Analytical performance of the primers and the ability of the signature to classify label-free patients with suspected sepsis into deterioration risk groups, defined by a worsening sequential organ failure assessment (SOFA) score and the need for ICU admission within the first 24 h, was determined in an independent cohort by routine semi-quantitative real-time reverse transcriptase polymerase chain reaction (RT-PCR, N=248). We then developed the molecular assay to perform quantitative one-step reverse transcriptase digital droplet PCR (ddPCR) detection of the RNA-based classifier at the POC using our LOC centrifugal microfluidic (CMF) system.^23,25,34^, A compact prototype instrument, the **<u>PRE</u>**cision me**<u>DI</u>**cine for **<u>C</u>**ri**<u>T</u>**ical care (**PREDICT**) device, that detects the Sepset classifier in less than 3 h using 50 µL of whole blood in the near-patient-environment was developed. Finally, we used PREDICT to classify patients into clinical deterioration risk groups using an independent cohort of label-free patients with suspected sepsis (N=30).

## Results

A high-level overview of the risk prediction classifier, platform development path and deployment workflow are shown in **Figure 1**. Three groups of independent cohorts (**Supplemental Table S1**) were used to reduce our original 99-gene signature down to 6 genes. We then validated, in a blinded fashion, the classifier’s ability to accurately identify amongst 248 well characterized patients, those who went on to deteriorate clinically, as defined by worsening of the sequential organ failure assessment (SOFA >=2) score and the need for ICU admission, 24 h post-initial assessment (early for patients with prospective sepsis).

**Figure 1:**
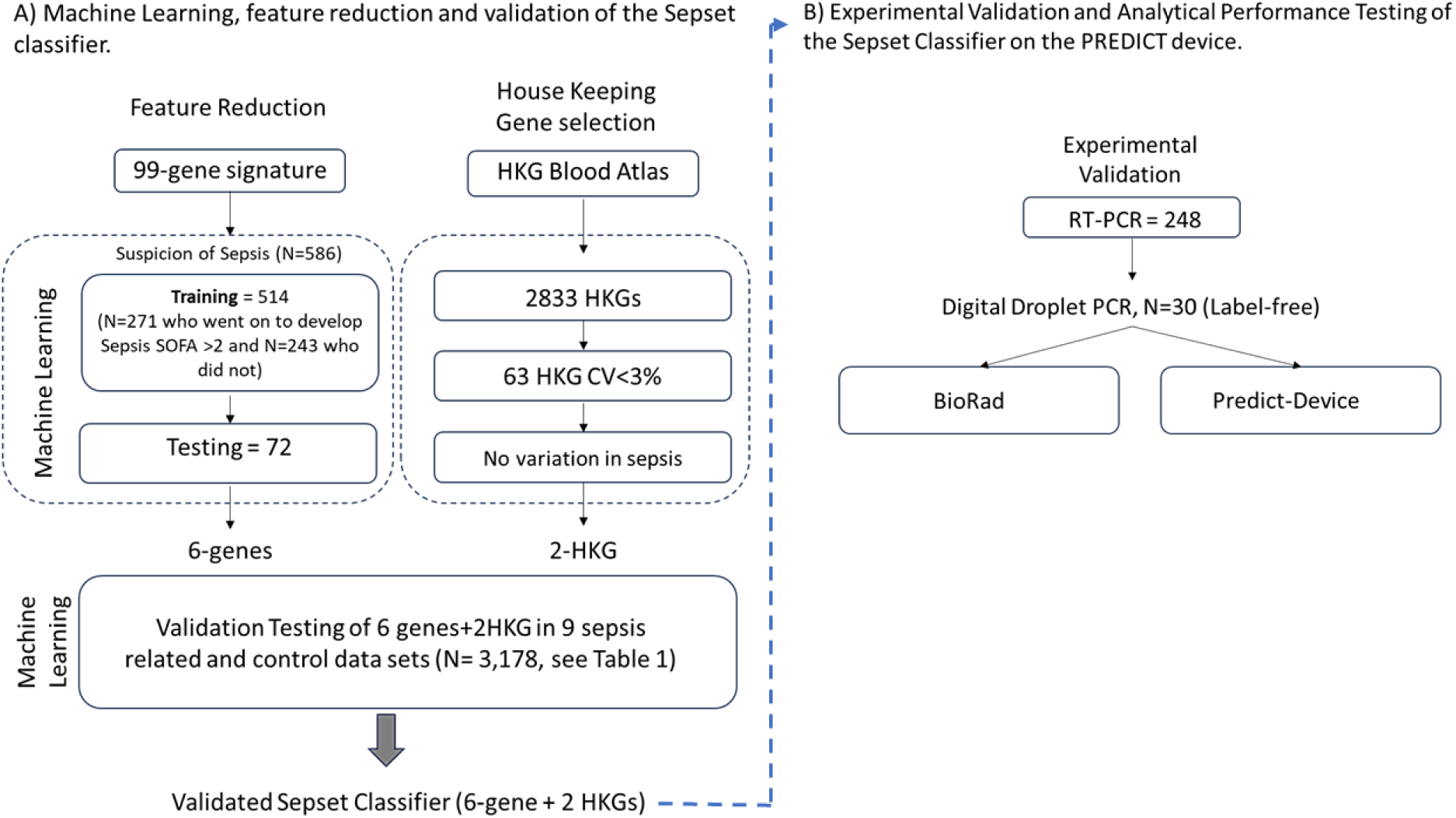
Overview of prediction classifier development, validation and analytical performance testing.

Our own whole blood RNA sequencing (RNA seq) data from 586 samples from 514 individuals (176 ICU and 338 ER patients); including 392 previously published^30^ and 194 new patient samples, were used to refine the gene expression signature able to predict clinical deterioration in patients with suspected sepsis.^30,31,35^, Briefly, these data were generated from whole blood tubes collected from consenting adult patients (> 18 years of age) with ethics approval, who presented with prospective sepsis, within the first 2 h of emergency room (ER) or within 24 h of intensive care unit (ICU) admission (see Methods) as previously published. ^30,31,35^

Each of the previously published 99 cellular reprogramming signature genes^30^ were tested for their ability to discriminate between 271 patients with suspected infections who went on to record a SOFA score >2 in the first 24 h of presentation and the 243 who did not. We used 514 first samples (out of total of 586) for machine learning (ML) model training and the remaining 72 second samples (out of 586) from the same 514 individuals as well as samples from 50 healthy individuals as validation samples. We selected six upregulated genes with the highest specificity, fold-change (FC) and lowest adjusted p-value (adj p-value, **Supplemental Table 2**). Increased FCs were associated with eventual worsening of SOFA scores as demonstrated in **Supplemental Figure S1A** showing the relative expression values for each of the putative signature genes as a function of the 24-hr SOFA score. This revealed a relationship between the relative gene expression of each signature gene and SOFA score (p<0.01 to p<0.0001). **Supplemental Figure S2B**, shows no relationship between lactate level and increased SOFA scores in sepsis samples (SOFA >= 2) vs. non-sepsis samples (SOFA < 2). Scattering of expression values around the mean implied that no single gene was discriminatory on its own, justifying the need for a multi-gene risk-prediction expression signature. The six-gene signature arose from the larger immune dysfunction (cellular reprogramming) signature.^31^ Genes comprising the signature and their abbreviated putative biological functions are shown in **Supplemental Table 3**.

To ensure we did not need to include positive and negative controls in the prognostic assay, the expression of each of our 6 genes was compared to two housekeeping genes (HKG) selected from the list of the 2,833 HKGs expressed in blood cells.^36^ Their expression variance (coefficient of variation, CV) was analyzed in the entire data set and 63 genes showing CV<3% across all of our datasets were selected as candidate HKG. These were filtered for lack of variance according to the clinical metadata considering sex, age, SOFA score, location (ICU vs ER), sepsis mortality, prediction and endotype^30^, as well as lack of variance in cancer patients (**Tables 1 and S1**). The accuracy of prediction using ratios of individual genes to HKG was tested. While no single gene expression ratio can discriminate those patients that are imminently at risk of clinical deterioration from those that are not, the collective set of 6 genes, when compared to 2 HKGs, gave excellent performance (**Table 1**).

**Table 1:**
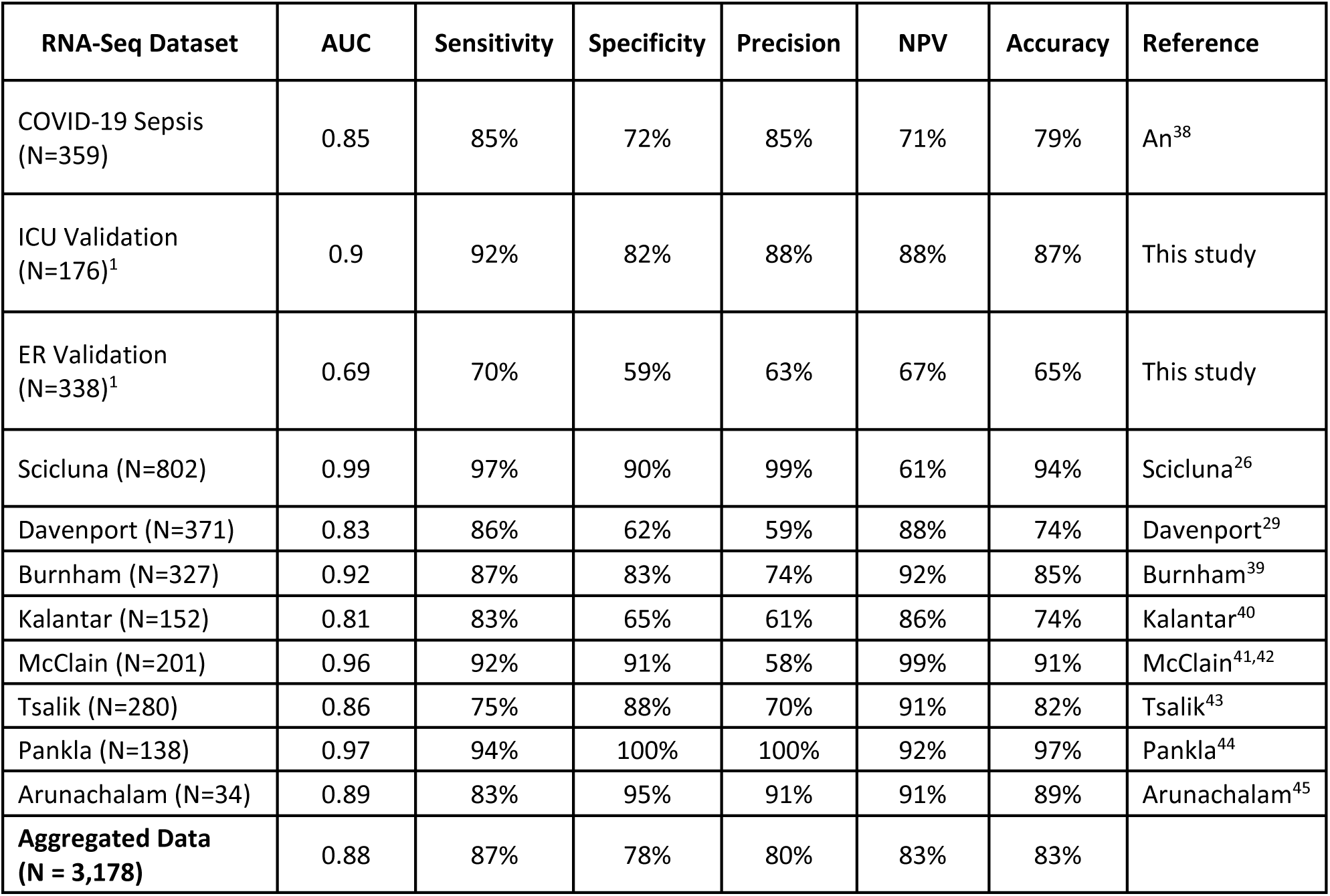
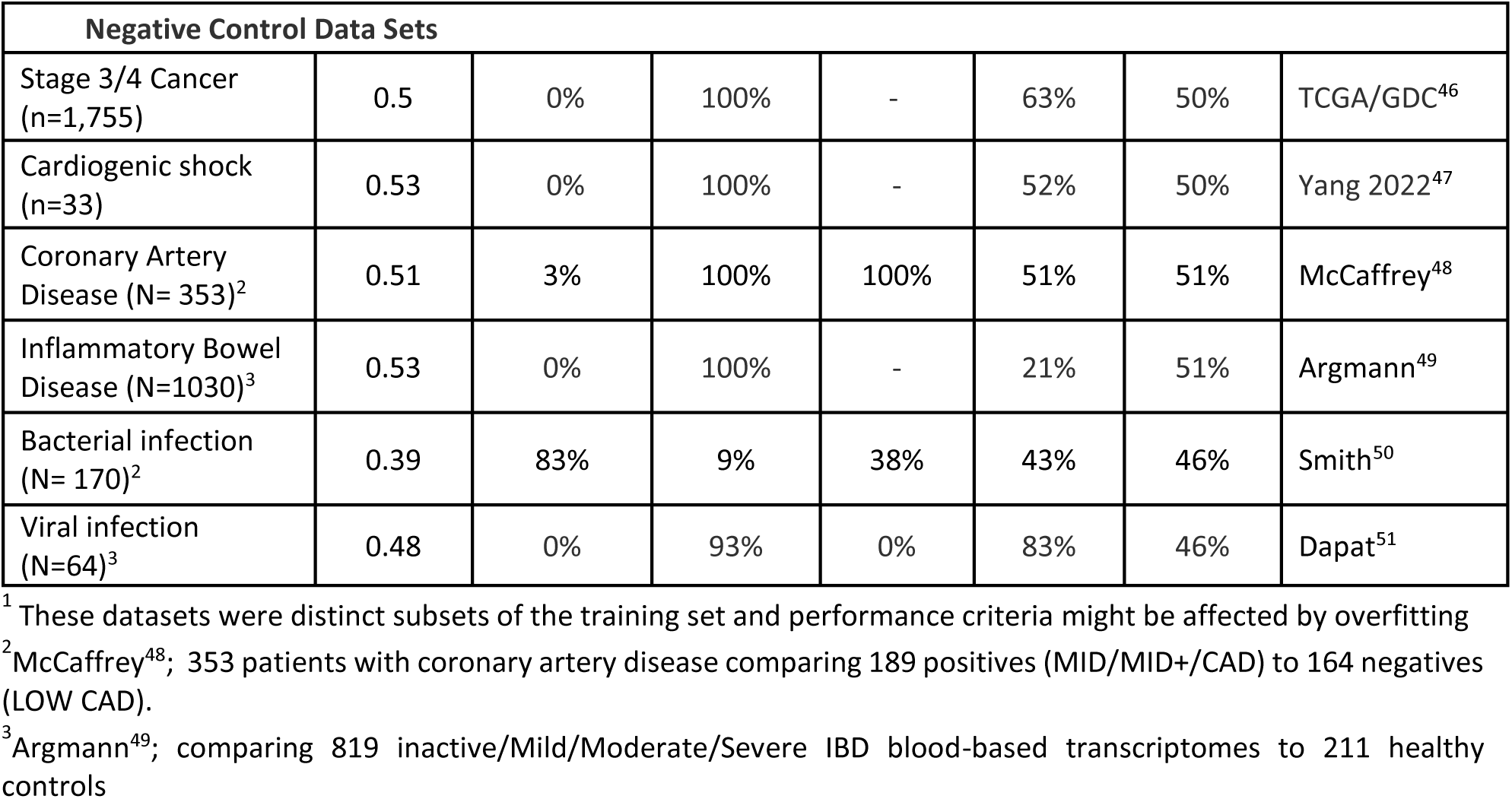
Summary Performance for Sepset signature and classifier. Performance of the Sepset signature was evaluated in different validation and testing datasets comprised of transcriptomic analyses of samples collected from various regions and settings. Aggregated results include analysis of patient samples subjected to transcriptomic analysis and available in the literature (see specific references). The internal data sets, samples indicated as “This Study”, were from an independent subset of patients from the Biomarkers of Lung Injury study (ClinicalTrials.gov ID NCT04747782), a prospective, observational, clinical study that collected whole blood from patients who presented to the ICU or ward with acute respiratory distress and suspected sepsis. Definitions: NPV = Negative Predictive Value; ICU, intensive care unit; ER emergency room.

To determine the best ML model to predict clinical deterioration within the first 24 h following clinical presentation, the input transcriptomic and associated clinical outcome data were analyzed using an ensemble of ML approaches. In patients with suspected sepsis, a SOFA cut-off score of 2 was used to discriminate predicted sepsis from non-sepsis.^1^ Using a change of SOFA score of >2 did not substantially change the results. Groups of patients were analyzed in a binary fashion (prospective sepsis cf. non-sepsis), using 18 machine learning algorithms, initially adopting a 10X cross-validation strategy. We chose to proceed with eXtreme Gradient Boosting (XGBoost)^37^, a regularizing gradient boosting framework/library involving matrices of decision trees, since it gave us the best performance (**Table 1 and Supplemental Figure S2**). Negative control data sets were used to reduce the likelihood of identifying false positives – patients with non-septic pro-inflammatory, shock, malignancy and infectious (bacterial or virus non-septic) conditions.

### A six-gene signature classified patients with sepsis in independent cohorts and was robust over time and a range of disease severity

We determined the ability of the six-gene compared to two-HKGs signature to accurately classify patients into pre-specified sepsis groups using nine sepsis-related gene expression datasets (N=3,178 (all samples) – 514 (training, or ICU + ER) = 2,664) downloaded from the Gene Expression Omnibus (GEO) repository. In these independent external cohorts, the reduced signature was able to discriminate between septic versus non-septic patients (Sepsis-3 criteria^1^), with a sensitivity ranging from 83-97% and balanced accuracy of 74-94%, a combined mean sensitivity of 85%, a specificity of 76%, and an accuracy of 81% (**Table 1**). We then analyzed samples from patients that were seen in the ER with suspicion of sepsis (N=338) versus those admitted directly to the ICU from the training cohort (N=176); this yielded sensitivity assessments of 70% and 92% respectively.^30^ To determine if the signature was robust over time and a range of disease severity, we assessed its performance in a validation cohort from the Biobanque Québécoise de la COVID-19 (BQC-19; Quebec COVID-19 Biobank) comprised of 359 blood samples taken at various times from 133 patients hospitalized with COVID-sepsis and exhibiting a range of severity.^33,38^ The ability of the reduced signature to discriminate between samples from patients that went on to clinically deteriorate (increased SOFA >2 or ICU admission) within the first 24 h from initial assessment, compared to those that did not was very good, with sensitivity and balanced accuracy of 85% and 79%, respectively. The set of six genes and two HKGs comprising the signature, we called here **Sepset**, was able to discriminate patients with suspicion of sepsis at risk of clinical deterioration.

### Sepset RNA amplification using RT-PCR reliably identifies patients that clinically deteriorated 24 h after initial assessment

To validate changes in gene expression using RT-PCR, we designed sets of PCR primers that spanned intron-exon boundaries (i.e., only contained in RNA not DNA), that amplified specific regions for each of the mRNA isoforms selected by the ML analysis (>90% of mRNAs), avoiding common single nucleotide polymorphisms (SNPs), for each of the 6 signature and 2 HKGs. Control experiments demonstrated that our primers successfully amplified mRNAs of interest extracted from human blood in an RNA copy number dependent manner, amplified as few as 10-20 copies of the gene and gave Ct curves in <25 cycles with blood mRNA. The PCR primers were then combined in triplex reactions and tested using whole blood RNA extracts from healthy donors. Reaction efficiencies were 100% with as little as 0.2 ng of input RNA (**Supplemental Figure S3**). We then tested the Sepset signature in a blinded fashion, using label-free whole blood samples from our in-house cohort of patients with prospective sepsis not included in the training set. As shown in **Table 1**, in 248 patients, Sepset was able to identify patients who went on to deteriorate with worsening SOFA score (fulfilling sepsis diagnosis based on SOFA ≥2 and suspicion of infection) within the first 24 h after sampling for RT-qPCR analysis, to those who did not. In these independent samples, Sepset demonstrated an AUC of 0.88 and a sensitivity of 94%.

### Association between Sepset with clinical features of clinical deterioration 24 h after initial assessment

In our 238 patients, we looked at the association between presence of Sepset with demographic characteristics, clinical features of sepsis, severity, and outcome measures at baseline, at 24 h (day 1) and 72 h (day 3) post-initial assessment (**Supplemental Table 4**). In the first 24 h, the signature was correlated with the number of comorbidities, SOFA score, the use of antibiotics, mechanical ventilation, and fluid resuscitation, but not oxygen or vasopressor therapy. Interestingly, we found no association with specific clinical parameters of organ dysfunction until day 3 (72 h) post-initial assessment. At 72 h, culture positivity and 28-day mortality correlated with the presence of the signature. At both 24 h and 72 h post-initial assessment, the signature was associated with important clinical outcome parameters such as the need for ICU admission, initiation of mechanical ventilation, use of fluid resuscitation, and initiation of antibiotics, as well as outcome parameters such as ICU length of stay. Unfortunately, we did not have data on dialysis requirements, but while presence of urine output at baseline was not associated, an increase in serum creatinine correlated with Sepset expression.

### Digital droplet PCR detected the Sepset classifier in 0.5 ng of patient-derived whole blood RNA

To migrate precision RNA-mark detection onto the LOC device, we designed the microfluidic platform to measure RNA expression using ddPCR. Herein, primers that spanned exon-exon junctions, were designed for each of the 8 genes (6 Sepset and 2 HKG) in the signature. The performance of each primer was benchmarked using the BioRad AutoDG/QX200 reader. Serial dilutions of human cDNA plasmids obtained from the mammalian genome collection^52^ were used to determine limits of detection (1 copy/μl) and dynamic range of the probes for each of the genes of interest (**Supplemental Figure S4A** left panel). Verification of primer performance was conducted using universal human cDNA (**Supplemental Figure S4A** middle panel) and 500 pg of patient cDNA (**Supplemental Figure S4A** right panels). A total of 500 pg of patient cDNA resulted in optimized partitioning of the positive and negative droplets and quantitation of the Sepset signature.

We then established the conditions for multiplexing using FAM and HEX labeled probes at final concentration of 5 μM (**Supplemental Figure S4B** left panel). Representative 2D tracing (**Supplemental Figure S4B** bottom right panel) showed partitioning of double negative (bottom left quadrant), FAM only (upper left quadrant), HEX only (lower right quadrant) and double positive (upper right quadrant) droplets. Probes alone (no cDNA) as a control rendered only double negative droplets (bottom right panel). Counts (in copies/µL) for each gene were obtained in triplicate and used to further refine the training of the predictive algorithm. Primers and probes used for ddPCR are presented in **Supplemental Table S5**.

### PREDICT platform detected the Sepset classifier at the point-of-need

Having validated the ability of the Sepset classifier to identify patients with ‘prospective’ sepsis, i.e. patients that will go on to clinically deteriorate within the first 24 h of clinical presentation, we sought to design and build an innovative POC device to perform the risk prediction at the bedside. Our previously published microfluidic technology^23,34^ was adapted to perform the molecular assay required to quantitate specific mRNAs using ddPCR at POC. The PREDICT system was developed as a stand-alone instrument that integrates the entire automated workflow (**Figure 2A**) for detection of the Sepset classifier directly from whole blood – without any pre-processing requirement. The analytical process was split between two microfluidic cartridges that when connected enable RNA isolation and ddPCR biomarker detection (see Supplemental materials for details including video 1 and video 2) to be performed in sequence automatically. Microfluidic cartridges are operated on a centrifugal platform (**Figure 2B**).^25,53,54^ Pneumatically-induced pressure imbalances allowed for transfer, valving, flow switching, inward pumping, and on-demand bubble-based mixing. The combination of centrifugation and pneumatic actuation rendered manipulation of liquids independent of wetting properties, which allowed for automation of the analytical protocol. The sample preparation cartridge (**Figure 2C**) contained buffers and reagents (**Supplemental Table S6**) to isolate RNA from whole blood, which was then used as a template for the ddPCR assay. The detection cartridge was adapted for monodisperse droplet generation and visualization (**Figure 2D–G**, **Supplemental Table S7**).

**Figure 2:**
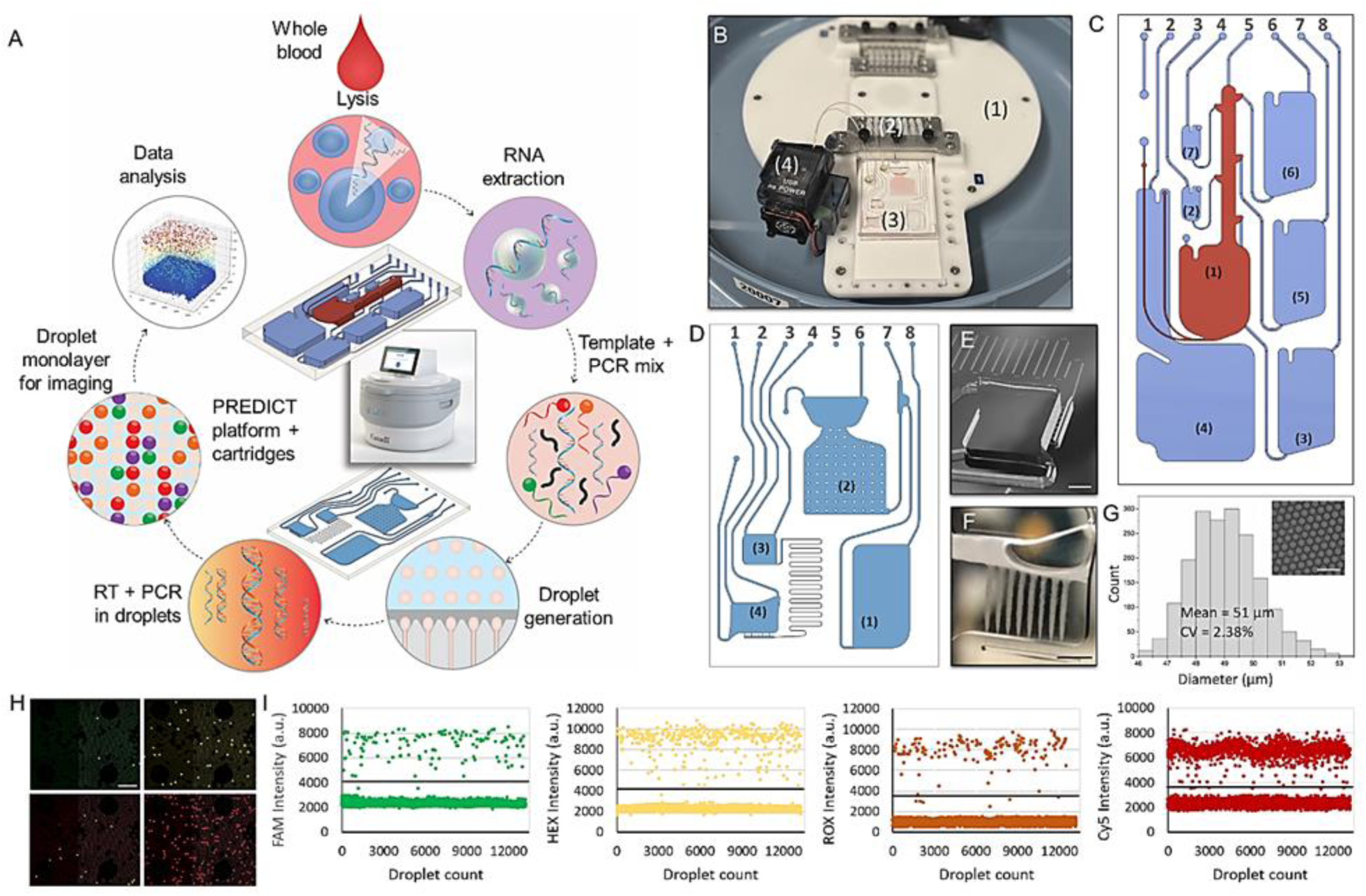
Microfluidic platform and cartridges. A, Microfluidic workflow for automation of RNA extraction and downstream ddPCR on the PREDICT platform and related cartridges. B, PREDICT platform showing rotor (1), pneumatic manifold (2), cartridge installed on the rotor (3) and connected to the external PCR tube using world-to-chip tubing inserted into the wirelessly controlled platform heater (4). C, RNA extraction cartridge design. D, Image of the ddPCR cartridge design. E, Micrograph showing close-up view of droplet generation chamber and nozzles. The scale bar is 2.4 mm. F, Micrograph showing close-up view of droplet streams generated using the ddPCR cartridge. The scale bar is 2.4 mm. G Size distribution of droplet diameter. The inset shows optical micrograph of droplet monolayer using ddPCR cartridge. The scale bar is 150 µm. H Example of acquired fluorescence images showing droplet monolayer within a region of the imaging chamber. For clarity, only a zoomed-in portion of the imaging chamber region is shown to increase visibility of droplets. The scale bar is 400 µm. I Intensity maps for different fluorophores. Horizontal lines denote the threshold for positive and negative counts.

The analytical workflow was conducted through a timed sequence of centrifugation and pneumatic actuation steps for which operational parameters are provided in **Supplemental Tables S8 and S9**. **Supplemental Figure S5** illustrates the sample preparation and detection cartridges during different stages of the process. The PREDICT system used a miniature epi-fluorescence imaging module for readout of droplets which comprises single-color excitation LEDs and proprietary optical filters for detecting probes labelled with FAM, HEX, ROX and Cy5 fluorophores. The instrument was adapted for recording fluorescence micrographs (**Figure 2H**) for each channel in sequence using an embedded monochrome CCD camera equipped with a 2× objective that had a sufficiently large field of view (e.g., 8 mm × 8 mm) to capture more than 10,000 droplets per image. The proprietary PREDICT software performed image analysis and sample quantification using droplet scatter intensity map for each fluorophore (**Figure 2I**) and ‘definetherain’ thresholding algorithm.^55,56^ Comparison of the performance of the PREDICT and BioRad QX200 ddPCR is presented in **Supplemental Figure S6**. A proprietary algorithm considered Sepset and HKG expression values and provided results as a probability of clinical deterioration within the next 24h.

We tested and validated the performance of the PREDICT prototype using label-free real-world samples from clinically annotated patients with prospective sepsis (N=30, Research Ethics Board # 20-078). These analyses were performed in a blinded fashion using a subset of the 248 technically and clinically benchmarked patients. The PREDICT system was able to predict clinical deterioration at presentation, as determined by worsening of SOFA score or need for ICU admission, providing a risk assessment to physicians regarding the consequent need for initiation of appropriate therapeutic (antibiotic, anti-viral, immune modulatory and/or monoclonal antibody) or supportive (ICU entry, mechanical ventilation, fluids) measures post-presentation. The sensitivity was 92%, specificity 89%, and overall accuracy 88% (**Table 2**), consistent with the RT-qPCR assays described above.

**Table 2:** Label-free performance validation of Sepset classifier. The PREDICT system was able to predict clinical deterioration at presentation, as determined by worsening of SOFA score or need for ICU admission compared to RT-qPCR and ddPCR. Definitions: RT-qPCR, reverse transcription quantitative polymerase chain reaction; ddPCR, droplet digital RT-PCR.

| Table 2: Label-free Validation of Sepset Classifier |  |  |  |  |  |  |  |
| --- | --- | --- | --- | --- | --- | --- | --- |
| RNA-Seq Dataset | AUC | Sensitivity | Specificity | Precision | NPV | Accuracy | Cohorts |
| RT-qPCR (N=248) | 0.88 | 94% | 72% | 84% | 88% | 83% | This study |
| ddPCR (N=30) | 0.91 | 100% | 83% | 80% | 100% | 92% | This study |
| PREDICT (N=30) | 0.88 | 92% | 83% | 79% | 94% | 88% | This study |

## Discussion

We have demonstrated the successful creation of a predictive sepsis gene expression signature and its migration to a novel POC platform PREDICT. This proof-of-concept demonstrates the feasibility of developing a rapid POC device that will enable physicians to conduct a prospective sepsis risk assessment at the point-of-need (in as little as 1-2 h after clinical presentation). In patients with suspicion of/or early sepsis, a positive result indicates a high probability that the patient will clinically deteriorate in the next 24 h (worsening SOFA score >=2 points and need for ICU admission), guiding physicians as to the need for ICU entry (in ER patients) and interventions like mechanical ventilation. Currently, the device provides a risk assessment in <3 h, well below the 6 ‘golden h’ recommended by the surviving sepsis campaign guidelines.^12^ In patients with suspected sepsis, PREDICT efficiently performs the entire workflow of RNA signature-based classifier detection, from sample-to-answer using just 50 µL of whole blood, without requiring pre-preparation of sample, and once transfer of eluate from the first to the second cartridge is automated, with no involvement from the clinical laboratory. Using real-world label-free patient samples, we showed the device has a high sensitivity of 92%, specificity of 89%, and an overall accuracy of 88% to identify the risk of imminent clinical deterioration in previously benchmarked patients with known outcomes.

There are two other established immune signature-based platforms, but neither appear to be intended for use like the Sepset signature. Septicyte RAPID test^57^ discriminates sepsis from non-infectious systemic inflammation and the original SeptiCyte^27^. This test, requires prior sample preparation, including RNA extraction from blood, separates patients into 4 bands with varying accuracy for prediction of sepsis cf. sudden SIRS, with only 55.8% being assigned to the highest and lowest bands and AUCs of 0.81-0.85^57^. In comparative studies, using Sepsis-3 definitions (96% of tested patients had sepsis), the AUC was 0.81, below that for all cohorts with the Sepset signature, except for ER patients not considered in the Septicyte study. Larger (29-31 genes) signatures that predict bacterial vs. viral infections (IMX-BVN3) and sepsis mortality (IMX-SEV), have been devised and again separate patients into 3 or 4 bands^58^. A recent performance study^58,59^ showed that the IMX-BVN3 signature adjudicated that, amongst patients with confirmed bacterial infections (64% of patients), 31% were possible and 6% were unlikely. Critically, however it is clear that both bacteria and specific viruses like COVID-19 can cause mechanistically almost-identical sepsis, and a positive cultures is only found in around 50% of patients^60,61^ even when using the latest molecular detection methods. Thus, discriminating bacterial from viral infections at the host response level may be useful with regards to indicating treatment options but not for identifying sepsis or identifying patients with suspicion of sepsis at risk of clinical deterioration. Conversely the sepsis mortality signature IMX-SEV^58^ shows a good AUC of 0.81 amongst ICU patients but this is separate from the Sepset signature that does not detect eventual mortality (**Supplemental Table S4**), in contrast to a separate signature we devised^33^. Overall, Sepset provides a simple binary risk assessment with high accuracy in very large numbers of patients (**Table 1**).

In our clinical study, the lower sensitivity of the test in ER patients with suspected sepsis possibly reflects the potential for early antibiotics to prevent development of organ dysfunction (74% received antibiotics Supplemental Table S4).^62^ We found no statistically significant association of Sepset prediction with presentation systolic or diastolic blood pressure, heart rate, respiratory rate, or temperature, but a modest association with altered mental status (as defined by the Glasgow Coma Scale Score <14, p=0.00099) and quick SOFA (qSOFA, p=0.013). We found the predictive function of the Sepset signature used in the PREDICT device is independent of initial clinical markers of severity (e.g., presentation lactate), and the association with parameters of deterioration was sustained for up to day 3 (72 h post-presentation) in ICU-admitted patients (p<0.005). These findings strongly suggest the Sepset classifier is not simply detecting a time window of severity; instead, it identifies a specific mechanism-based signature. This signature provides information about immunological cell reprogramming which impairs immune cells’ ability to respond to pathogens. This impairment is associated with future clinical deterioration, independent from initial clinical markers of severity. In keeping with this hypothesis, we found that the addition of other clinical or laboratory information, outside of the Sepset classifier, is not required for its performance in predicting which patients are likely to go on deteriorate over the next 24 h. This means that, other than suspicion of sepsis, no other information should be needed for accurate prediction of the clinical course over the first 24 h following initial assessment. We have prospectively performed validation of this finding in the COVID cohort that was studied longitudinally,^38^ which suggests that, the PREDICT device may be used in low resource or remote settings, by non-expert personnel, to make immediate risk assessments about level of care.

Currently, there are no universally accepted diagnostic or prediction tools routinely used at the bedside for patients with suspected sepsis; especially during the critical early stages of disease where the risk of clinical deterioration may not be clear. Despite early adequate treatment, many patients fail to improve; highlighting the heterogeneity of the syndrome and the need for pathogen-agnostic risk stratification as well as novel therapeutics.^63,64,65^ The condition remains misdiagnosed in approximately 30% of patients.^66^ Poor specificity of current syndromic diagnostic approaches results in unnecessary use of drugs (e.g., antibiotics and steroids) and exposure to harm (iatrogenicity) associated with hospital and ICU-level care. They also fail to predict, for those patients who are admitted to the ward or are sent home, whether and/or how patients deteriorate and need higher levels of care,^67^ placing a considerable financial and human burden on both hospitals and patients, directly affecting prognosis. The primary purpose of the SOFA score is to objectively describe organ (dys)function rather than to predict outcome, so no associated equation developed for prediction is currently in use.^68^ There is evidence from a range of observational studies however, that even a small change in SOFA score is associated with a persistent trend in mortality.^69^ Recently, Seymour et al.^70^ investigated the validity of the SOFA score to predict in-hospital mortality in patients with suspected infections and found it to have an AUC-ROC of 0.74. Therefore, predicting what the SOFA score will be 24 h post-presentation may have important clinical value.

Unfortunately, there are important limitations to the calculation and utility of the SOFA score. For example, it requires input from healthcare personnel, and samples need to be processed in a clinical laboratory which may prove challenging when access to clinicians/nurses and/or the laboratory is limited. While a qSOFA has been proposed as a potential solution in low resource settings, there are some intrinsic problems with its utility in guiding early management and qSOFA show poor accuracy (42-46% in 2 meta analyses^71,72^). One of the advantages of PREDICT is its potential to provide information about the risk of clinical deterioration independent of the initial SOFA/qSOFA.

The advantages of Sepset and PREDICT include no pre-preparation of the sample, requirement of very small volumes of blood and the accuracy afforded by the use of ddPCR. Importantly, PREDICT is intended for prediction of risk of clinical deterioration in patients suspected of sepsis, so this technology differs fundamentally from most other available tests. Other studies have used retrospective electronic transcriptional data to predict 30-day mortality in adults.^28^ The advantage of using a small-sized gene classifier, and the ability to use highly accurate multiplex PCR, needs to be balanced with the need for precision, although we have observed no diminution of effectiveness of the 6 gene signature compared to the 99-gene cellular reprogramming signature from which it was devised.^31^

An important limitation of our study is that the PREDICT and Sepset classifier are yet to be tested prospectively to determine if its implementation would lead to improved management and outcomes. Although we were able to show high overall accuracy in studies comprising 2,559 patients, there is a paucity of existing cohorts with high-quality (especially RNA-Seq) transcriptomic data for validation, despite the high frequency of sepsis (49M cases per year). Very few studies have collected early sepsis and/or longitudinal data with accompanying biological samples linked to granular metadata. Since the pandemic, the importance of collecting clinically annotated biological samples for translational research has significantly improved. While showing that we can predict clinical deterioration in patients with bacterial and COVID-19 sepsis, the study should be expanded to other viral causes of sepsis and to important subsets of bacterial sepsis (e.g., caused by particular bacteria). Importantly, we did not have information regarding longer-term outcomes, such as 90-day and hospital mortality for the entire cohort. Our data comes from observational studies, so it precludes us from determining whether the current classifier is informative regarding response to therapy. In contrast, the classifier can be tested in any other cohort even if similar clinical data have not been collected, as long as clear indicators of clinical deterioration, such as a SOFA scores are available. Given that the PREDICT-based test is conducted in the immediate near-patient environment, the expectation is that there will be no need for storage, transport, or handling of tubes, simplifying its use at the bedside.

In summary, the current study describes the development of a molecular risk classifier for clinical deterioration and onset of sepsis, and a novel POC device to measure this at the bedside, as well as the proof-of-concept demonstration using real-world patient samples. The important feature of the classifier and the technology is that no additional information, other than suspicion of sepsis, is required to obtain a risk assessment that can be used in a clinically actionable fashion at the bedside. Moreover, in the future, no expert personnel or equipment are required to prepare the sample or interpret the results. Prospective testing of the device and the classifier will be fundamental in moving forward to determine the clinical utility of the tool. However, the technology can be adapted to measure virtually any nucleic acid-based marker, making it modular, and therefore adaptable to easily measure both existing and emerging markers. This adaptability could potentially enhance the predictive performance of the classifier.

## Methods

### Ethics

This is a multicenter, secondary analysis of a prospectively recruited longitudinal cohort study enrolling consecutive patients with suspected sepsis. All patients were enrolled under local ethical board approval. Informed written consent was obtained upon enrollment from the patient or their legal representative. The Clinical Research Ethics Board (REB) of the University of British Columbia (UBC) provided ethics approval for all sequencing and bioinformatics studies, carried out in a manner blinded to patient identity (approval number REB#H20-02441, REB#H17-01208). Patients recruited and enrolled at Unity Health Toronto were included in accordance with protocol approved by the St. Michael’s Hospital ethics board (REB#: 20-078). Patients’ data were extracted from the in-hospital electronic medical records, de-identified, and assigned random identification numbers which were used throughout the project. All experiments performed at the NRC involving human samples were approved by the NRC’s Ethics Board (NRC REB 2021-57) and experiments were performed according to NRC’s policies governing human subjects that follow applicable research guidelines compliant with the laws in the province of Québec.

### Sample collection, RNA isolation and cDNA conversion

Patient samples were collected in Pax Gene tubes and total RNA was isolated using standard protocol for Qiagen RNAeasy mini kit. RNA was assessed first using NanoDrop One spectrophotometer (Thermo Scientific) and A260/A280 values were between 1.8 and 2.2, with typical yields in the range of 6–8 µg total.

RNA Integrity Number (RIN) was determined using the Agilent 2100 Bioanalyzer (Agilent Technologies). Following the standard Nanochip protocol, samples with RIN values >7.0 were used for conversion to cDNA. Input volumes for reverse transcription were calculated using the concentration from the bioanalyzer (approximately 500 ng total was used per sample) and a High Capacity cDNA Reverse Transcription Kit (Applied Biosystems) was used following standard protocols.

### Discovery dataset

The whole transcriptome (RNA-seq) data from 586 whole blood samples from different countries and continents comprised our patient cohort. 514 samples were collected and used for discovery analyses (i.e., the discovery cohort). The remaining 72 samples were secondary samples collected from 72 individuals, which were excluded from discovery analyses (to prevent same-individual artifacts) and used as a validation cohort. The sepsis severity associated with the discovery cohort (514) was based on the SOFA score of the patient at 24 h after the first sample collection: 271 samples with SOFA ≥ 2 were sepsis, and 243 samples with SOFA <2 were non-sepsis.

### Sepsis signature and housekeeping gene selection

We tested 99 cellular reprogramming (CR) genes as potential sepsis markers^31^. We used DESeq2 to perform differential gene expression analyses and chose the genes that had the highest up-regulation (positive fold-change) in high severity samples. We also estimated the predictive accuracy of each CR gene by setting the sensitivity to 75%. We picked six genes (RETN, S100A8, MCEMP1, S100A12, CYP1B1 and HK3) that had the best results in both analyses for the SepsetER model.

We analyzed 2,833 housekeeping genes^36^ in our discovery cohort of 514 samples to set a baseline for RNA quantity and sequencing depth. We selected housekeeping genes (HKG) with high and consistent expression across all samples based on mean and variance. We then examined the expression variance of the top 20 HKG candidates across key clinical factors such as age group, gender, sepsis severity, patient location, mortality, etc. The two housekeeping genes (PTP4A2 and CHTOP) with the lowest variances were used to set a baseline for the SepsetER model.

### ML algorithm construction and testing

Our own RNA-Seq data from 873 patient samples^30^, was used for feature (gene) reduction using ML. An additional 1241 transcriptomes from patients were used for testing the derived signature. Three major groups of datasets were used for biomarker development – training, validation and testing. The discovery data set (N=586) was first tested by 10X cross validation and randomly divided into a <u>training</u> (90% of the samples) used for the construction of the models (10,000+ models) and a <u>test</u> dataset (10% of the samples) to assess the best model (**Supplemental Figure S2**). We trained 18 different machine learning algorithms on the transcriptomic profiles of the 514 discovery cohort samples. The algorithms were: K-Nearest Neighbors (KNN), Ridge Regression (RR), Lasso regression (LR), Elastic Net (EN), Partial Least Square (PLS), Linear Discriminant Analysis (LDA), Regularized Discriminant Analysis (RDA), Quadratic Discriminant Analysis (QDA), Bayesian Generalized Linear Model (BL), Naïve Bayes (NB), Support Vector Machines (SVM), Decision Tree (DT), Random Forest (RF), Adaptive Boosting (AB), Stochastic Gradient Boosting Model (GBM), Extreme Gradient Boosting (XGB), Neural Network (NN), and Multilayer Perceptron (MLP). We tested each model with different parameters and chose the best one based on the AUC-ROC using 10-fold cross validation, repeated 10 times. We then validated the performance of each model with the additional 72 validation samples (that were not in the training dataset).

We tested the SepsetER model with multiple methods. We used various sepsis transcriptome datasets (from microarray and RNA-seq platforms) with over 3,000 sepsis and healthy samples to evaluate the SepsetER sepsis classification model. We also trained other published sepsis gene-signatures with our training dataset and compared them with SepsetER. The SepsetER model, using the Extreme Gradient Boosting (XGB) algorithm, performed better than all other signatures, with a median AUC-ROC of 0.85 in all testing datasets.

### Design of primers and probes

The expression of 6 top genes of interest were assessed based on the selection of the highest fold changes with respect to severity of disease in the ICU cohort. These genes are HK3 (108bp), RETN (78bp), S100A12 (122bp), S100A8 (122bp), MCEMP1 (131bp), CYP1B1 (114bp). The housekeeping genes were also selected based on stable expression: PTP4A2 (138bp) and CHTOP (113bp).

Primers (IDT) for these genes have been designed to span the exon-exon junction to avoid amplification of genomic DNA and to cover the different isoforms. The amplicons’ sizes range between 78bp and 138bp. The probes were synthesized (IDT) with either FAM, HEX, ROX or Cy5 fluorescent labels and a ZEN/3’ Iowa Black FQ (IABkFQ) double quencher when possible to reduce background noise.

Two fourplex reactions were designed to include 3 genes of interest and 1 HKG for normalization. As such one reaction targeted: CYP1B1, MCEMP1, S100A12 and PTP4A2, and the other: HK3, RETN, S100A8 and CHTOP. The sequences of the primers and probes are provided and described in the supplementary S.

Specificity and sensitivity of the primers and probes were first assessed by performing qPCR standard curves of the individual targeted genes from human universal cDNA (P/N 637223, Clontech/TaKaRa Bio) and comparing the efficiency with the multiplex reaction. The results obtained were then used to design the multiplex ddPCR reactions in order to ensure appropriate amplification of differentially expressed genes and avoid amplification bias.

### Commercial duplex ddPCR

For optimal results, recommendations made in the Droplet Digital PCR Applications Guide (bulletin 6407) were followed. We used equal concentrations of cDNA for droplet generation following the protocol for ddPCR supermix for probes (bio-rad.com/web/ddPCRsmxProbes). Briefly, a 20 µL reaction set-up consists of 2X supermix, 20X probes (a duplex of FAM and HEX), equal concentrations of the patient samples (500 pg), and RNase-free water. The bulk solution (in a 96-well plate) is applied to the AutoDG (automated droplet generator) where the solution is partitioned into 10,000 individual water-in-oil droplets. The 96 well plate is foil sealed and put into the C1000 thermal cycler (BioRad) where the individual droplets are subjected to the following conditions: 10 min at 95 °C, 40 cycles of 30 s at 94 °C and 1 min at 60 °C, followed by 10 min at 98 °C and a 4 °C hold. Subsequently, the droplets were read in the QX200 Droplet Reader using FAM and HEX channel readout in the QuantaSoft software. After data acquisition, the QC of the samples was assessed (ensuring equal droplet numbers generated) and samples were selected in the well selector tool under the Analyze tab. Samples were all manually thresholded using the values from probe alone readout and confirmed in 2D tracings of the duplexed reaction. Samples were tested in duplicate. The concentration reported is “copies/ng DNA” of the final 1X ddPCR reaction.

## Microfluidic device fabrication

### Sample preparation cartridge

Microfluidic channels and reservoirs were carved into a block (50 mm × 100 mm × 6 mm) of Zeonor 1060R (Zeon Chemicals) using precision machining (Q350 CNC Mill; Menig Automation). The machined polymer piece was cleaned with isopropanol (Sigma-Aldrich) and dried with a stream of nitrogen gas. The microfluidic circuit was sealed using adhesive film (ARclear 93495, 40 μm in thickness; Adhesive Research) applied on a polycarbonate sheet (250 μm in thickness; McMaster-Carr).

### Detection cartridge

The microfluidic circuit was fabricated in polydimethylsiloxane (Sylgard 184; Dow Corning) using replica molding. A multi-level SU-8/silicon master mold was made by sequential photolithographic patterning of multiple layers (10, 30 and 50 µm in thickness) of SU-8 photoresist (GM1060 and GM1070; Gersteltec) spin-coated onto a 6″ silicon wafer (Silicon Quest International) in conjunction with flood exposure at 365 nm (Hg i-line) through a chrome/quartz glass photomask (Photronics) using an EVG 6200 mask alignment system (EV Group). SU-8 resist was developed in propylene glycol monomethyl ether acetate (Sigma-Aldrich) for several minutes, followed by rinsing with isopropanol (Anachemia) and drying with a stream of nitrogen gas. Bake steps were performed on a programmable hot plate (HS40A; Torrey Pines Scientific) using recommended time and temperature settings. The liquid pre-polymers of PDMS were mixed at a ratio of 10:1 (w/w) elastomer base/curing agent, poured onto the SU-8/silicon master mold, and cured at 85 °C for 1 h. The cured PDMS replica was bonded to a glass substrate following oxygen plasma activation (HI RF power, 900 mTorr for 30 s; Harrick Plasma).

### Microfluidic assay implementation

#### Total RNA extraction from whole blood

Total RNA was extracted from 50 μL of whole blood collected in PAXgene tubes using custom Galenvs Total RNA kit (Galenvs) following manufacturer’s recommendations. Briefly, whole blood aliquot is mixed with 50 μL PBS and introduced onto the cartridge for automated protocol or processed manually for extraction in tubes. The mixture was first combined with 20 μL Proteinase K, and mixed. Lysis/binding buffer was then added to the solution and incubated at 55 °C for 10 min. For manual extraction in tubes, a DynaMag magnetic rack (Thermo Fisher Scientific) was used to capture magnetic nanoparticles. Following the capture of the RNA bound to the beads, two consecutive wash steps are performed. Elution was performed in 25 μL of nuclease-free water (Sigma-Aldrich). On-chip extraction of total RNA was performed using the automated protocol implemented on the centrifugal platform with the same reagents and volumes as for the manual extraction. For the on-chip capture of magnetic nanoparticles (MNPs), the external magnetic field was provided by a nickel-plated neodymium alloy disk magnet (D201, 1/8” in diameter, 1/32” in thickness; K&J Magnetics) which remained inserted in the designated area on the cartridge for the entire duration of the automated assay. The extracted RNA was subsequently used in downstream RT-qPCR for assessment of RNA extraction efficiency as well as in on-chip ddPCR for determination of transcript copy number.

#### qPCR

cDNA obtained from different patients were analyzed in a multiplexed qPCR using primer-probe sequences for genes of interest and housekeeping genes as internal controls for normalization. Each qPCR reaction consisted of 5 μL 10X PCR Buffer (Qiagen), 8 μL HotStar Taq Plus DNA Polymerase, 3 μL 25 mM MgCl_2_, 1 μL dNTPs, 5 μL 10X primer-probe mix (final concentration of 1 μM and 0.5 μM, respectively), 2 μL template, and 26 μL nuclease-free water (Sigma-Aldrich), for a total volume of 50 μL. Samples were tested in duplicate. A no-template control (NTC) reaction was included to assess for contamination. Thermal cycling was performed according to the manufacturer’s recommended protocol in a Bio-Rad CFX96 Touch Real-Time PCR Detection System (Bio-Rad). To quantify the copies of genes of interest each qPCR run included serial dilutions of cDNA (Takara) generating as such a standard curve. Cq values were plotted against the log concentration and linear regression was used to determine standard curves. The efficiency of each assay was 100 ± 10% and the *R*^2^ of each standard curve was >0.98.

#### RT-ddPCR

The ddPCR reaction master mix comprised of 5 μL 10X PCR Buffer (Qiagen), 8 μL HotStar Taq Plus DNA Polymerase, 8 μL 100X QuantiTect Virus RT Mix (Qiagen), 3 μL 25 mM MgCl_2_, 1 μL dNTPs, 5 μL 10X primer-probe mix (final concentration of 1 μM and 0.5 μM respectively), 2 μL template, and 18 μL nuclease-free water (Sigma-Aldrich), for a total volume of 50 μL. Template input was 2 µL of cDNA, RNA, or nuclease-free water for NTC samples. On-chip ddPCR assay was performed using an automated protocol implemented on the centrifugal platform. Briefly, droplets containing template input in ddPCR reaction master mix were generated on-chip in fluorinated carrier oil (5% 00-8 FluoroSurfactant in HFE7500) (RAN Biotechnologies). The resultant emulsion was then transferred to the platform heater and cycled following manufacturer’s recommended protocol (20 min at 50 °C, followed by 5 min at 95 °C and 40 cycles of 15 s at 95 °C and 45 s at 60°C, with ramp rate of 1 °C/s). Following thermal cycling, the emulsion was transferred to the chip for fluorescence imaging and data analysis. All experiments were performed in duplicate (no significant differences).

#### Implementation of the on-chip SepsetER classifier detection process

The automated RNA extraction protocol (**Supplemental Figure S5A**) starts with introduction of the sample in the RNA extraction chamber and installation of the cartridge on the platform. The software then executes a pre-programmed protocol sequence by initiating the platform to rotate. The first step of the automated workflow includes the transfer of a Proteinase K solution to the RNA extraction chamber, and bubble mixing. The lysis/binding buffer containing magnetic nanoparticles is subsequently transferred to the sample, mixed, and incubated for 10 min at 55 °C. The rotation speed is then increased to capture MNPs, and the lysate is transferred to the waste chamber. Two wash steps are then carried out sequentially by transferring the wash solutions from their respective chambers to the RNA extraction chamber. Finally, the purified RNA is eluted in the clean elution buffer.

To begin the cDNA synthesis and ddPCR protocol, a 2 µl aliquot of the eluted RNA is introduced on the ddPCR cartridge in the PCR mix chamber containing the RT-ddPCR master mix. Two cartridges, each having capacity to perform a single fourplex ddPCR reaction are operated in parallel to detect the 8-gene classifier. The automated sequence (illustrated in **Supplemental Figure S5B**) commences by transferring the fluorinated oil into the droplet imaging chamber, followed by emulsification of RT-ddPCR master mix in the droplet generation chamber. The latter is performed by applying a positive pressure onto the ports of the master mix chamber to push the liquid through the resistive serpentine channel entering the array of nozzles connected to the shallow terrace merging into a deep reservoir of the droplet generation chamber. Upon completion of the droplet generation process, the rotation speed is reduced and positive pressure is applied in order to gently transfer the emulsion off-chip into the PCR tube located on the platform heater, using the world-to-chip interface. Following thermal cycling, the emulsion is transferred back on chip by applying positive pressure. The droplets sitting on the top of the oil in the neck of the imaging chamber are subsequently arranged in a monolayer suitable for imaging by applying a low negative pressure at ports of the oil reservoir. This step withdraws (back) the fluorinated oil from the imaging chamber into the oil reservoir and gently lowers the droplets into the shallower portion of the chamber. The pressure is slowly decreased to 0 psi until the monolayer formation is complete and the rotor is stopped, thus allowing the acquisition of fluorescence images for subsequent analysis.

## Data Availability

Previously published data can be obtained from GEO. Data from internal validation cohort can be obtained upon request

## Acknowledgements

We acknowledge our membership in the Canadian Critical Care Trials (CCCTG), Canadian Critical Care Translational Biology Group (CCCTBG, CCDS) and Sepsis Canada (RH and CCDS) and are grateful for their insightful review of the manuscript (Drs. A. Fox-Robichaud, and Keith Walley). We are also indebted to Drs. Laurent Brochard, Arthur Slutsky and Gilbert Walker for the insightful comments. We gratefully acknowledge funding from the Collaborative R&D Initiative Pandemic Response Challenge Program Grant Application from the National Research Council of Canada to CCDS, RH, TV and LM (2020); the Canadian Institutes of Health Research (CIHR) Collaborative Health Research Projects (CHRP, Natural Sciences and Engineering Research Council of Canada partnered, FDN-433426 to CCDS, TV, LM, MG and DB); CIHR to CCDS (FDN-420463); CIHR to AB and CCDS (Emerging COVID-19 Research Gaps & Priorities, FDN-466806); CIHR COVID-19 Rapid Research Funding and CIHR FDN-154287 to RH. RH holds the University of British Columbia Killam Professorship and previously held a Canada Research Chair. CCDS holds the Robert and Dorothy Pitts Research Chair in Acute and Emergency Medicine and AB holds the Cara Phelan Chair of Critical Care Medicine at the University of Toronto. The COVID cohort studies were made possible through open sharing of data and samples from the Biobanque Québécoise COVID-19, funded by the Fonds de recherche du Québec – Santé, Génome Québec and the Public Health Agency of Canada, the University of Toronto COVID Biobank funded by the Canadian Foundation for Innovation (CFI); and the CFI John R. Evans Leaders Fund (CFI-JELF, 2020) and St Michael’s Hospital Foundation (2020) for the creation of the PREDICT-biobank at Unity Health Toronto.

## Supplemental Materials

**Supplemental Table S1:**
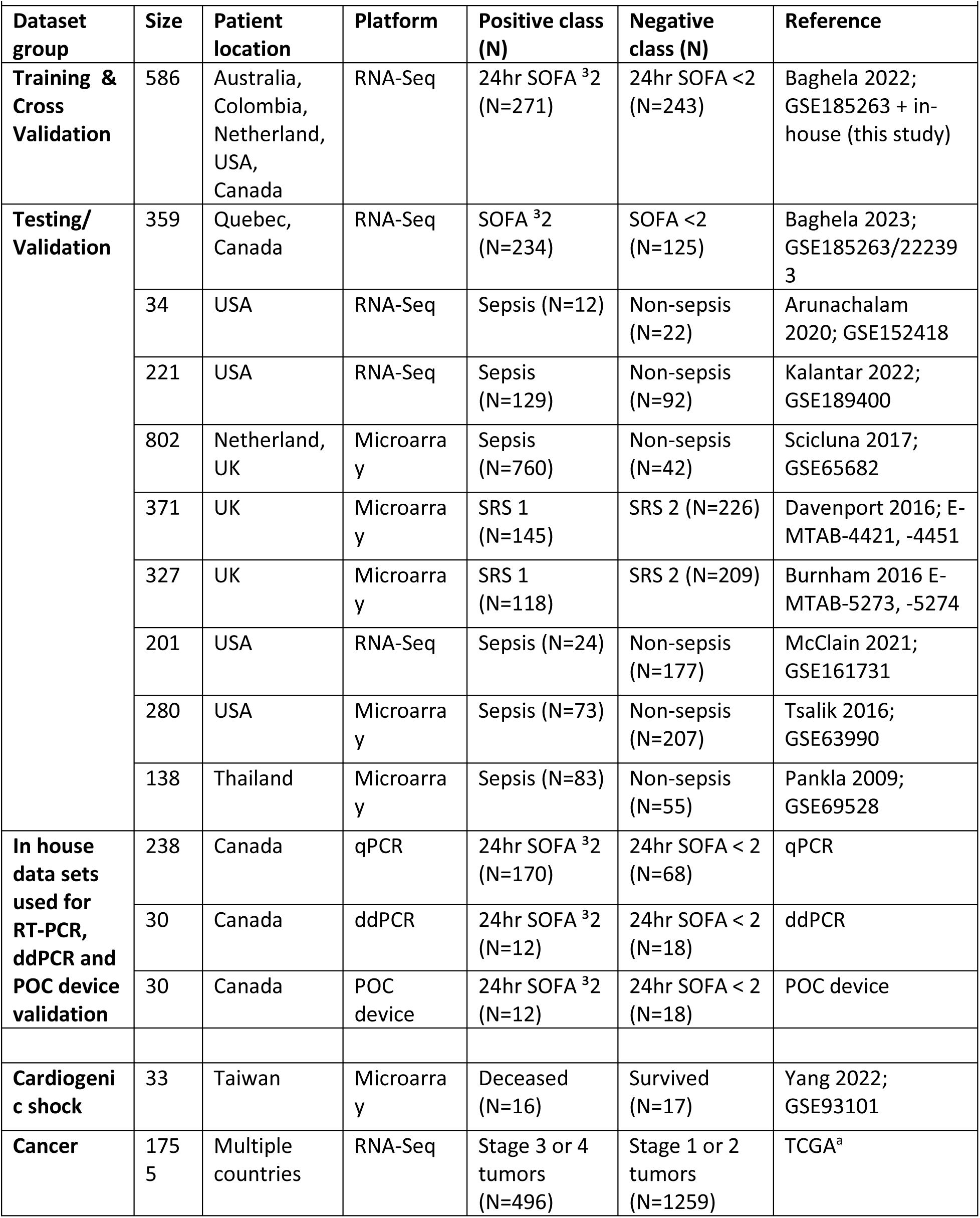
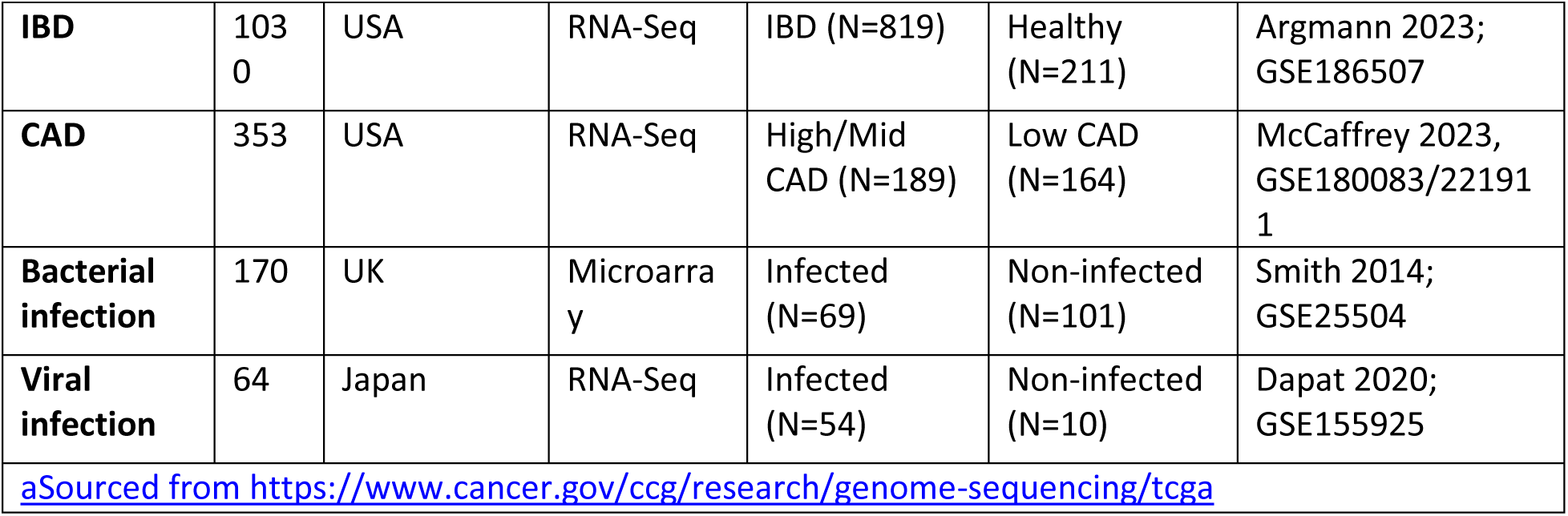
Data sets used for derivation, validation and testing of classifier.

|  |  |  |  |  |  |  |
| --- | --- | --- | --- | --- | --- | --- |
| Dataset group | Size | Patient location | Platform | Positive class (N) | Negative class (N) | Reference |
| **Training & Cross Validation** | 586 | Australia, Colombia, Netherland, USA, Canada | RNA-Seq | 24hr SOFA <sup>3</sup>2 (N=271) | 24hr SOFA <2 (N=243) | Baghela 2022; GSE185263 + in-house (this study) |
| **Testing/ Validation** | 359 | Quebec, Canada | RNA-Seq | SOFA <sup>3</sup>2 (N=234) | SOFA <2 (N=125) | Baghela 2023; GSE185263/222393 |
|  | 34 | USA | RNA-Seq | Sepsis (N=12) | Non-sepsis (N=22) | Arunachalam 2020; GSE152418 |
|  | 221 | USA | RNA-Seq | Sepsis (N=129) | Non-sepsis (N=92) | Kalantar 2022; GSE189400 |
|  | 802 | Netherland, UK | Microarray | Sepsis (N=760) | Non-sepsis (N=42) | Scicluna 2017; GSE65682 |
|  | 371 | UK | Microarray | SRS 1 (N=145) | SRS 2 (N=226) | Davenport 2016; E-MTAB-4421, -4451 |
|  | 327 | UK | Microarray | SRS 1 (N=118) | SRS 2 (N=209) | Burnham 2016 E-MTAB-5273, -5274 |
|  | 201 | USA | RNA-Seq | Sepsis (N=24) | Non-sepsis (N=177) | McClain 2021; GSE161731 |
|  | 280 | USA | Microarray | Sepsis (N=73) | Non-sepsis (N=207) | Tsalik 2016; GSE63990 |
|  | 138 | Thailand | Microarray | Sepsis (N=83) | Non-sepsis (N=55) | Pankla 2009; GSE69528 |
| **In house data sets used for RT-PCR, ddPCR and POC device validation** | 238 | Canada | qPCR | 24hr SOFA <sup>3</sup>2 (N=170) | 24hr SOFA < 2 (N=68) | qPCR |
|  | 30 | Canada | ddPCR | 24hr SOFA <sup>3</sup>2 (N=12) | 24hr SOFA < 2 (N=18) | ddPCR |
|  | 30 | Canada | POC device | 24hr SOFA <sup>3</sup>2 (N=12) | 24hr SOFA < 2 (N=18) | POC device |
| **Cardiogenic shock** | 33 | Taiwan | Microarray | Deceased (N=16) | Survived (N=17) | Yang 2022; GSE93101 |
| **Cancer** | 1755 | Multiple countries | RNA-Seq | Stage 3 or 4 tumors (N=496) | Stage 1 or 2 tumors (N=1259) | TCGA<sup>a</sup> |
| **IBD** | 1030 | USA | RNA-Seq | IBD (N=819) | Healthy (N=211) | Argmann 2023; GSE186507 |
| **CAD** | 353 | USA | RNA-Seq | High/Mid CAD (N=189) | Low CAD (N=164) | McCaffrey 2023, GSE180083/221911 |
| **Bacterial infection** | 170 | UK | Microarray | Infected (N=69) | Non-infected (N=101) | Smith 2014; GSE25504 |
| **Viral infection** | 64 | Japan | RNA-Seq | Infected (N=54) | Non-infected (N=10) | Dapat 2020; GSE155925 |
| aSourced from https://www.cancer.gov/ccg/research/genome-sequencing/tcga |  |  |  |  |  |  |

**Supplemental Table S2:** Selection of genes for the Sepset signature. Presented are analyses performed per gene on 443 patient samples comprising 213 with SOFA scores ≤ 2 and 230 with SOFA scores ≥2 at 24 hours post-presentation. Shown are the final selection of 6 genes based on specificity, fold change and adjusted p-values (padj). Increased FCs were associated with eventual worsening SOFA score (≥5). All genes were previously reported as part of the 99-gene cellular reprogramming/endotoxin tolerance signature.

| HGNC symbol | Fold Change<br>SOFA $\geq 2$ | Specificity | P-adjusted | Fold Change<br>SOFA $\geq 5$ |
| --- | --- | --- | --- | --- |
| RETN | 4.28 | 0.77 | 1.46E-13 | 11.39 |
| MCEMP1 | 2.23 | 0.69 | 1.40E-05 | 5.50 |
| HK3 | 2.16 | 0.69 | 1.45E-09 | 3.77 |
| S100A8 | 2.49 | 0.65 | 5.76E-09 | 6.57 |
| CYP1B1 | 2.08 | 0.64 | 4.39E-08 | 2.03 |
| S100A12 | 2.49 | 0.63 | 5.01E-07 | 6.01 |

**Supplemental Table S3:** Putative function of genes in the Sepset signature: The HUGO Gene Nomenclature Committee (HGNC) official gene symbol. Gene name and putative function in GeneCards the Human Gene database and joint functional prediction in Enrichr, adjusted p-value and odds ratio describe likelihood that functional enrichment prediction using REACTOME or GO Molecular function was identified by chance alone.

| HGNC symbol | Gene Name | Functional Prediction | Source |
| --- | --- | --- | --- |
| RETN | Resistin | Antibacterial activity against both Gram positive and Gram negative bacteria | https://www.genecards.org/cgi-bin/carddisp.pl?gene=RETN&keywords=MCEMP1 |
| MCEMP1 | Mast Cell Expressed Membrane Protein 1 | Involved in regulating innate immune responses | https://www.genecards.org/cgi-bin/carddisp.pl?gene=MCEMP1&keywords=MCEMP1 |
| HK3 | Hexokinase 3 | phosphorylate glucose to produce glucose-6-phosphate, the first step in most glucose metabolism pathways | https://www.genecards.org/cgi-bin/carddisp.pl?gene=HK3&keywords=HK3 |
| S100A8 | S100 Calcium Binding Protein A8 | Intracellular activity: facilitates leukocyte arachidonic acid trafficking and metabolism, modulation of tubulin-dependent cytoskeleton during migration of phagocytes and activation of the neutrophilic NADPH-oxidase. Extracellular: acts as an alarmin, antimicrobial, oxidant-scavenging and apoptosis-inducing activities | https://www.genecards.org/Search/Keyword?queryString=%20S100A8 |
| CYP1B1 | Cytochrome P450 Family 1 Subfamily B Member 1 | Monoxygenase which catalyze reactions involved in drug metabolism and synthesis of cholesterol, steroids and other lipids. | https://www.genecards.org/cgi-bin/carddisp.pl?gene=CYP1B1&keywords=CYP1B1 |
| S100A12 | S100 Calcium Binding Protein A12 | Acts as an alarmin, activity involves recruitment of leukocytes, promotion of cytokine and chemokine production, and regulation of leukocyte adhesion and migration. | https://www.genecards.org/cgi-bin/carddisp.pl?gene=S100A12&keywords=S100A12 |
| All 6 genes together (RETN, MCEMP1, HK3, S100A8, CYP1B1, S100A12) | Sepset genes | REACTOME Pathway: Neutrophil Degranulation R-HSA-6798695 (adjusted p= 0.0002310, odds ratio= 84.18)<br>GO Molecular Function: RAGE Receptor Binding GO:0050786 (adjusted p=0.00003777, odds ratio=1665.67) | https://maayanlab.cloud/Enrichr/enrichr |

**Supplemental Table S4:** Baseline demographic and clinical characteristics and correlation with clinical traits, interventions and outcomes at recorded at 24 and 72 h. Data were collected from consenting adult patients (> 18 years of age) with ethics approval, who presented with prospective sepsis, within the first two hours of emergency room (ER) or within 24 h of intensive care unit (ICU) admission at baseline, 24 h and 72 h. Abbreviations: ICU (intensive care unit), SOFA (sequential organ failure score), WBC (white blood cell count), P/F ratio (ratio of the partial pressure of oxygen over fraction of inspired oxygen), IQR (interquartile range). A significant correlation between features and Sepset is denoted by p≤0.05 using Χ^2^/Fisher exact test for categorical variables and analysis of variance for continuous variables as appropriate with Benjamini and Hochberg correction.

| **Demographics and Characteristics** | **Baseline Median IQR** | **24 hrs** | **72 hrs** |
| --- | --- | --- | --- |
| Age | 52.5 ± 22.75 | ns | 1.60E-05 |
| Sex, Female | 45.50% | ns | 2.66E-02 |
| Number of Comorbidities | 2 ± 4.75 | 4.50E-03 | 9.99E-09 |
| Location ICU | 31% | 5.90E-03 | 8.80E-05 |
| SOFA score | 4 ± 4.5 | 4.70E-05 | 3.00E-08 |
| Glasgo Coma Scale (GCS) | 13 ± 2 | ns | 2.90E-04 |
| Heart Rate (HR, beats per minute) | 82.5 ± 23.5 | ns | ns |
| Mean Arterial Pressure (mmHg) | 81 ± 20 | ns | ns |
| Temperature (Degrees Centigrade) | 36.2 ± 0.77 | ns | 1.10E-02 |
| Oxygen Saturation (Percent) | 96 ± 4.25 | ns | 4.90E-02 |
| Total Serum Bilirubin (μmol/L) | 10 ± 4.5 | ns | 9.70E-06 |
| WBC Count (10<sup>9</sup>/L) | 8.58 ± 7.92 | ns | 8.10E-11 |
| Platelets Count (10<sup>9</sup>/L) | 207 ± 127.5 | ns | 1.40E-03 |
| Serum Creatinine (μmol/L) | 68 ± 136.25 | ns | 3.30E-03 |
| P/F Ratio | 227 ± 118.5 | ns | ns |
| D-dimers (μg/L) | 1,198 ± 949 | ns | ns |
| C-Reactive Protein (mg/L) | 97.3 ± 157.4 | ns | ns |
| Serum Lactate (mmol/L) | 1.7 ± 1.4 | ns | 1.20E-03 |

| **Interventions and Outcome Measures** | **Baseline Median IQR** | **24 hrs** | **72 hrs** |
| --- | --- | --- | --- |
| Antibiotics given | 74% | 1.08E-02 | 8.39E-03 |
| MV initiated | 28% | 2.16E-02 | 4.71E-05 |
| Fluid resuscitation received | 84% | 4.10E-02 | 2.38E-02 |
| Oxygen Therapy received | 89% | ns | 2.20E-03 |
| Urine output at presentation | 92% | ns | ns |
| Inotropic Support (days) | 0 ± 7 | ns | ns |
| Length of ICU Stay (days) | 4 ± 9 | 5.90E-03 | 8.80E-05 |
| Hospital Length of Stay (days) | 9 ± 2 | 4.10E-09 | 4.10E-09 |
| Suspected site of infection Lung | 62% | ns | 1.60E-05 |
| Suspected site of infection Abdomen | 33% | ns | 5.77E-02 |
| Suspected site of infection CNS | 0% | ns | ns |
| Blood Culture Pathogen | 18% | ns | 5.89E-06 |
| 28-Day Mortality | 17% | ns | 2.62E-07 |
| 90-Day Mortality | 2% | ns | ns |

**Supplemental Table S5.**
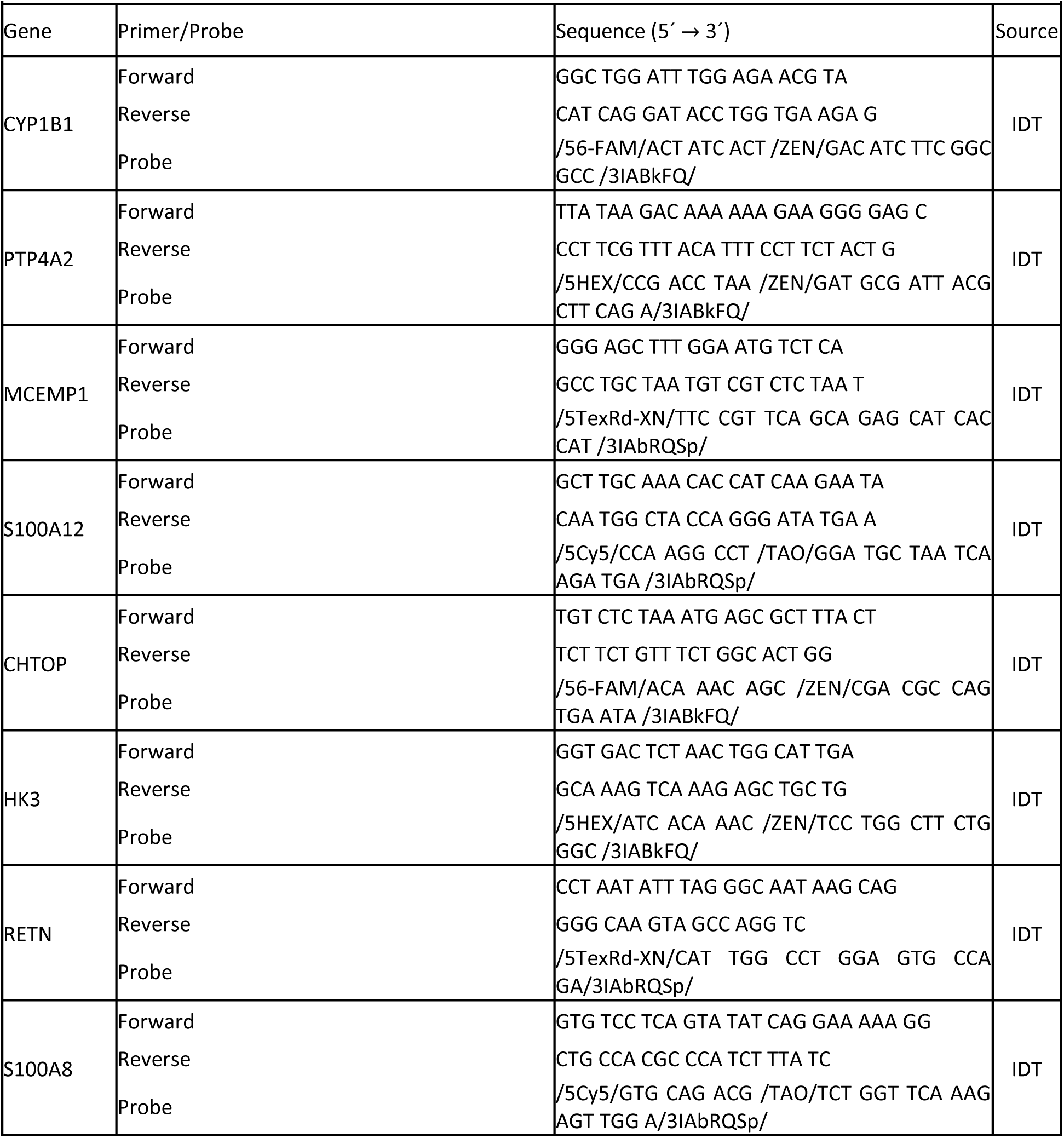
Primers and Probes used for ddPCR.

**Supplemental Table S6.** Characteristics of reservoirs implemented in the RNA extraction cartridge fluidic design and corresponding reagents.

|  |  |  |  |  |  |
| --- | --- | --- | --- | --- | --- |
| Reservoir # | Description | Location on cartridge | Capacity (μL) | Reagent | Reagent volume (μL) |
| 1 | RNA extraction | Top side | 980 | Sample: PBS (1:1) | 100 |
| 2 | Proteinase K | Bottom side | 70 | Proteinase K | 20 |
| 3 | Lysis/binding | Bottom side | 1,125 | Lysis/binding buffer | 400 |
| 4 | Waste | Bottom side | 3,500 | – | – |
| 5 | Wash 1 | Top side | 1,020 | Wash 1 buffer | 550 |
| 6 | Wash 2 | Top side | 1,020 | Wash 2 buffer | 550 |
| 7 | Elution | Top side | 70 | Elution buffer | 25 |

**Supplemental Table S7.**
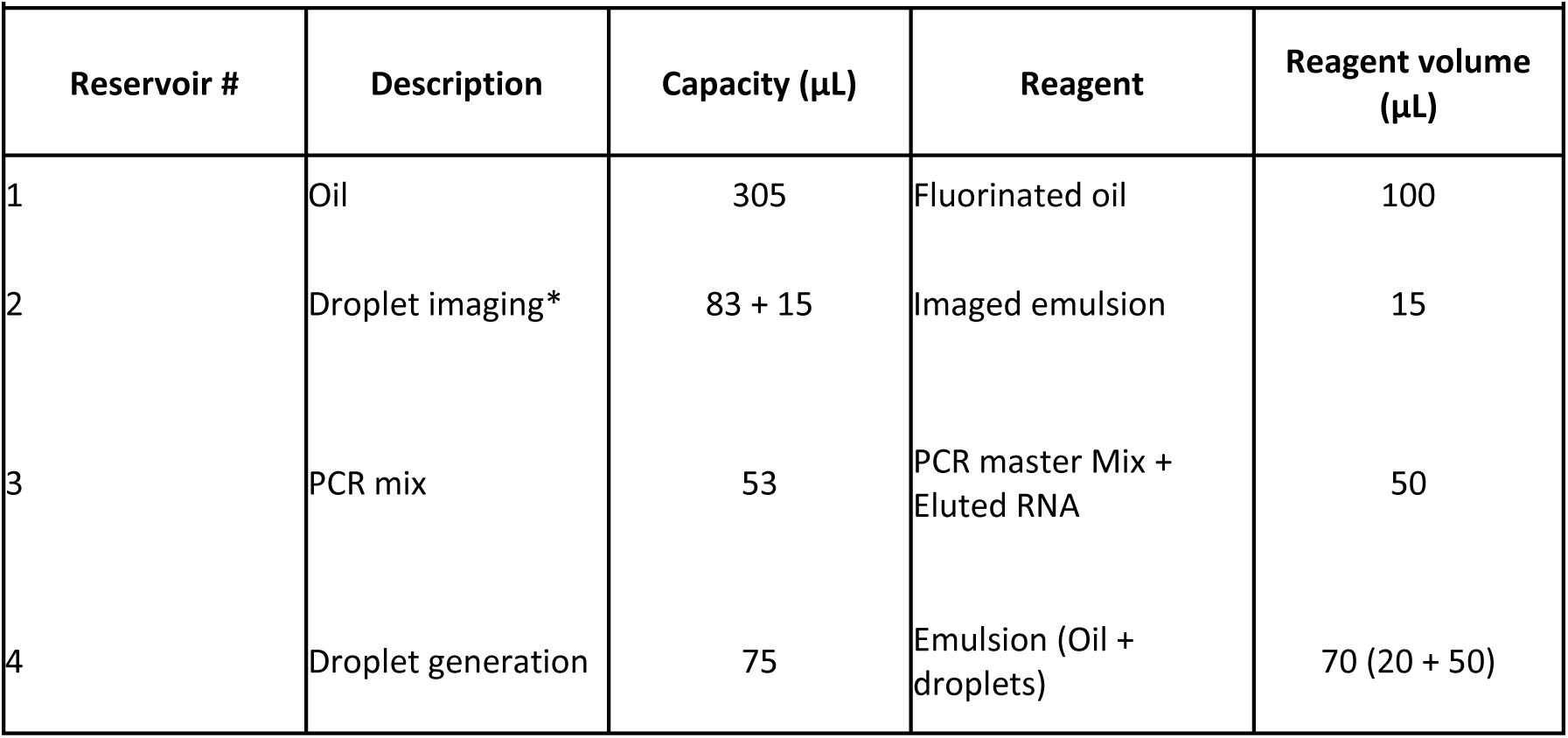
Characteristics of reservoirs implemented in the ddPCR cartridge fluidic design and corresponding reagents.

**Table S8.** Microfluidic implementation of the automated RNA extraction protocol.

| Step | Operation | Active ports (#) | Applied pressure (psi)<sup>a</sup> | Rotation speed (rpm) | Duration |
| --- | --- | --- | --- | --- | --- |
| 1a | Transfer of Proteinase K | 3 | 1 | 600 | 15 s |
| 1b | Bubble mixing of sample and Proteinase K | 2 | 1 | 600 | 0.5 s (10×) |
| 1c | Incubation | – | – | 400 | 1 min |
| 2a | Transfer of Binding buffer | 8 | 1 | 600 | 15 s |
| 2b | Bubble mixing of sample and MNPs | 2 | 1 | 600 | 0.5 s (10×) |
| 2c | Incubation at 55 C | – | – | 400 | 10 min |
| 2d | Capture of MNPs | – | – | 800 | 5 min |
| 3 | Transfer of lysate to waste chamber | 2 | –0.4 to –1.4 | 800 | 1 min |
| 4a | Transfer of wash buffer 1 | 7 | 2.5 | 500 | 15 s |
| 4b | Incubation | – | – | 400 | 5 min |
| 4c | Transfer of wash buffer 1 to waste chamber | 2 | –0.4 to –1.4 | 800 | 1 min |
| 5a | Transfer of wash 2 buffer | 6 | 2.5 | 500 | 20 s |
| 5b | Incubation | – | – | 400 | 2 min |
| 5c | Transfer of wash buffer 2 to waste chamber | 2 | –0.4 to –1.4 | 800 | 1 min |
| 5d | Air dry of MNP pellet | 2 | –1.5 | 800 | 1 min |
| 6a | Transfer of elution buffer | 4 | 1.9 | 700 | 5 s |
| 6b | Bubble mixing of elution buffer and MNPs | 2 | 1 | 700 | 0.2 s (30×) |
| 6c | Capture of MNPs | – | – | 800 | 5 min |
| 7 | Transfer of eluted RNA sample to an external tube | 1 | −1.5 | 600 | 30 s |

<sup>a</sup> Values are relative to atmospheric pressure.

**Supplemental Table S9.**
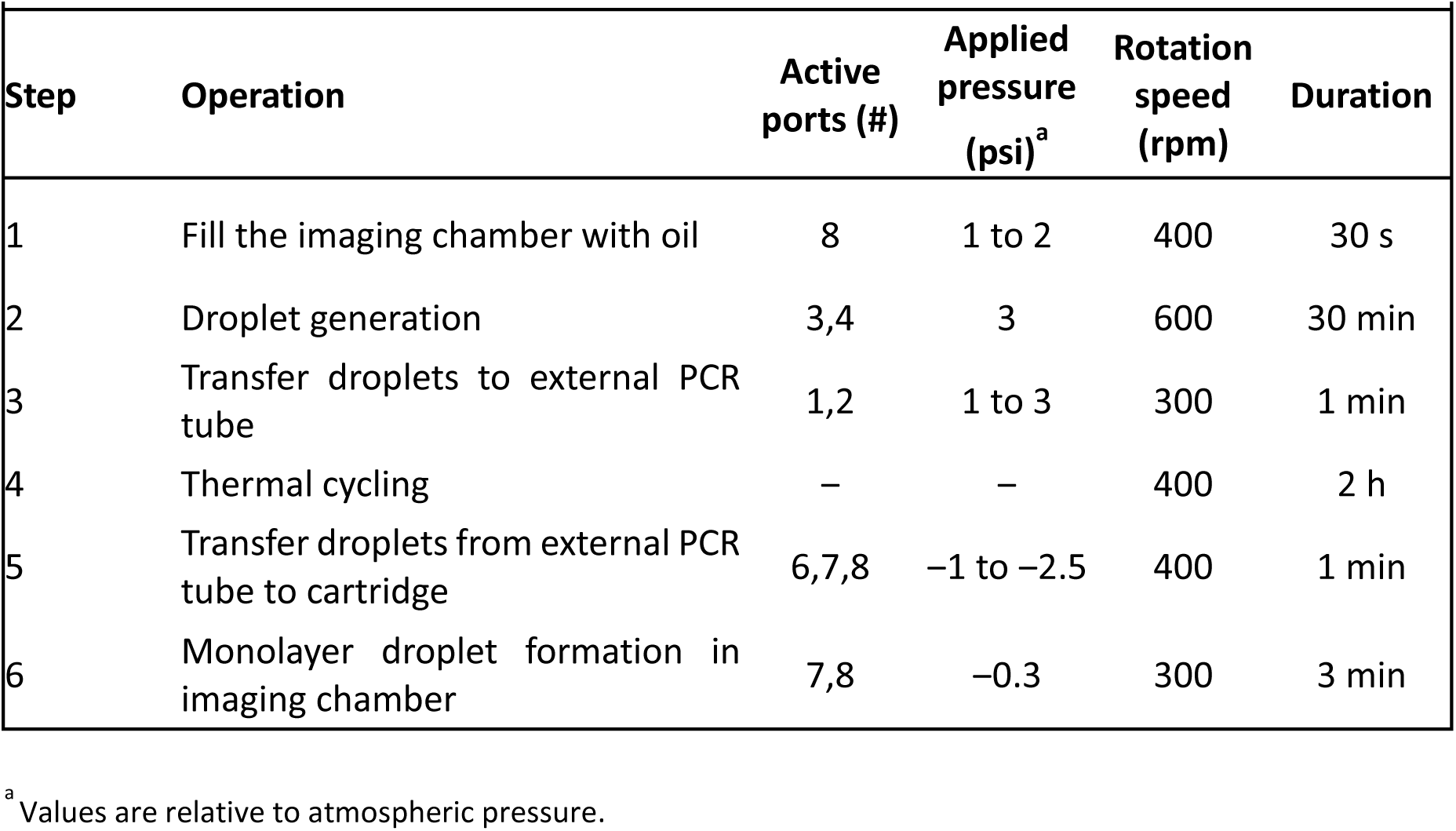
Microfluidic implementation of the automated ddPCR protocol.

**Supplemental Figure S1:**
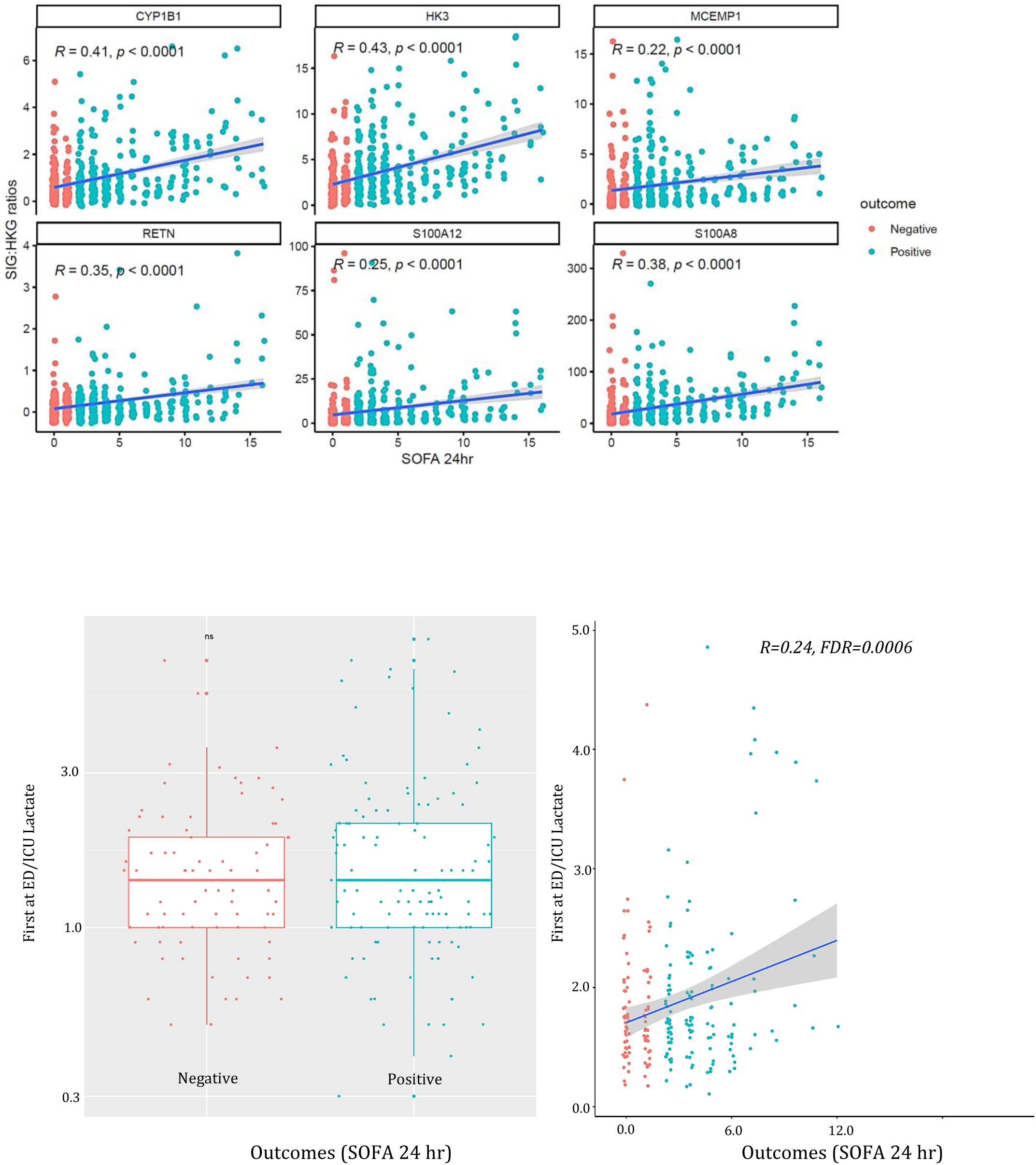
**A**) Relationship between 24-hour SOFA score and the relative gene expression of each of the Sepset-signature genes demonstrating a relative increase in expression as a function of 24-hour SOFA score. Red = Sepsis negative SOFA <2; Blue = sepsis positive SOFA ≥2. **B)** Relationship between baseline SOFA and serum lactate with clinical deterioration. Boxplots shows no difference between lactate level between sepsis samples (positive = SOFA >= 2) vs. non-sepsis samples (negative = SOFA < 2). The scatterplot shows poor correlation between lactate level vs. SOFA score (R = 0.24, suggesting lactate – as a marker of clinical severity – can only account for ∼5% variation in SOFA scores).

**Supplemental Figure S2.**
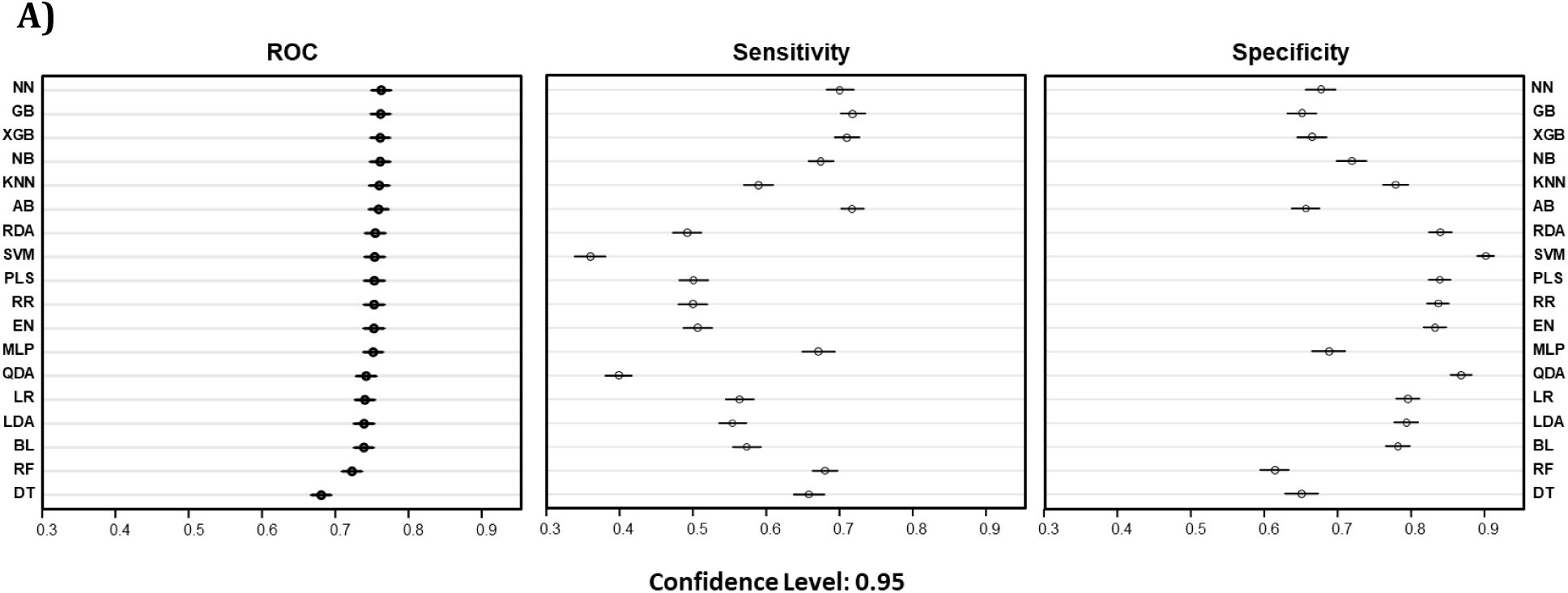
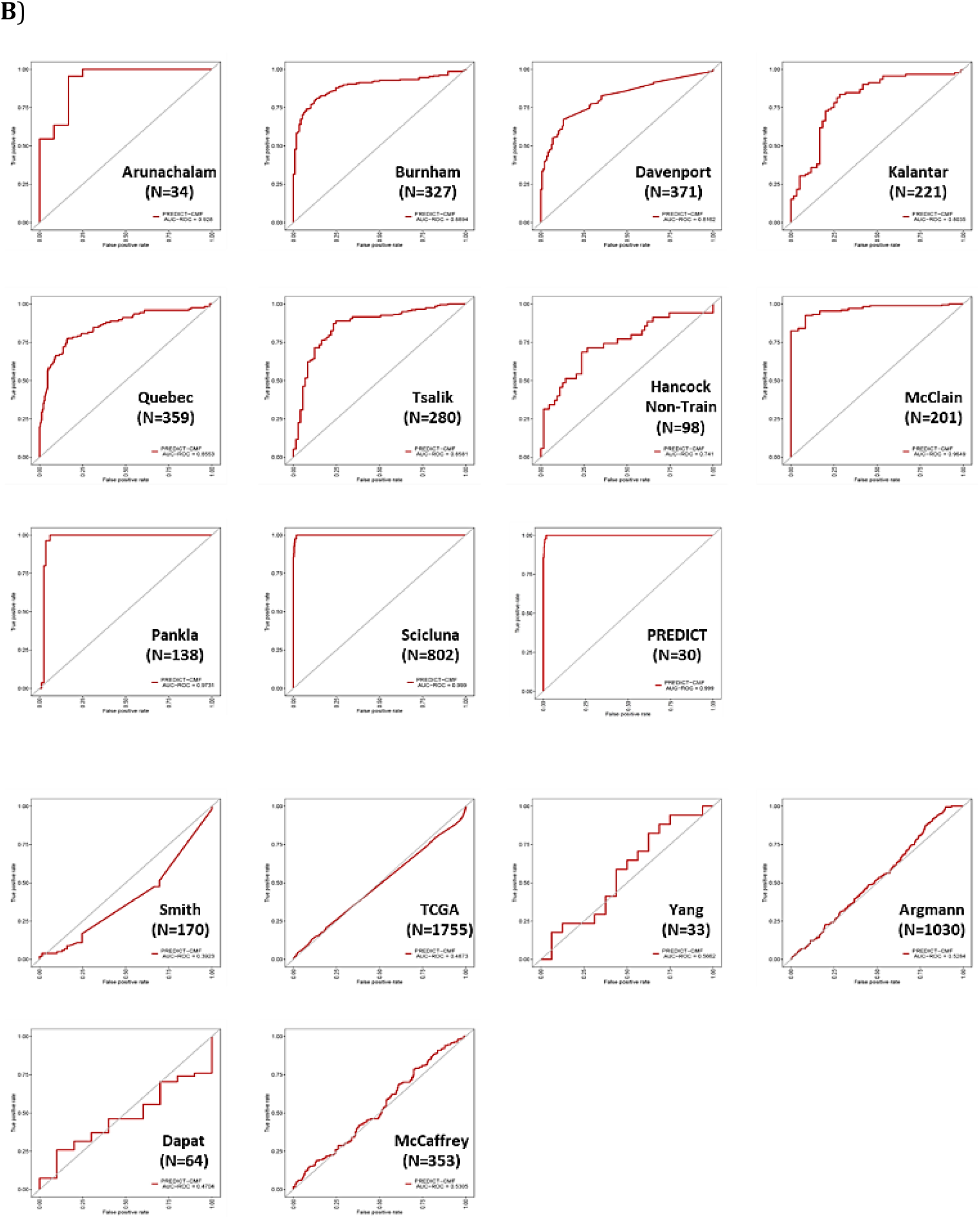
Selection of the Sepset (and 2 HKG) gene expression signature. Schematic of the patient cohort analyzed and the methodology of machine learning/algorithm development to predict clinical outcomes based on expression levels. We used an ensemble of single feature selection algorithms to identify the optimal biomarker gene set in terms of both predictive performance and robustness. **A**) AUC-ROC of Training data set. The ensemble approach combined results from thirteen different selectors: K-Nearest Neighbors (KNN), Support Vector Machine (SVM), Red Deer Algorithm (RDA), Gradient Boosting Machine (GBM), Neural Network (NN), Partial Least Squares regression (PLS), Lasso Regression (LASSO), Elastic net (ELASTIC), Radio Frequency (RF), Background subtraction (BG), Naïve Bayesian (NB), Linear Discriminant Analysis (LDA), and Flexible Gradient Boosting (GB and XGB). **B**) The AUROC for the ability of the Sepset classifier to correctly classify label-free samples from data-sets downloaded from GEO (not used in the derivation) with known group assignment (septic vs non-septic as defined by suspicion of sepsis + SOFA>2).

**Supplemental Figure S3.**
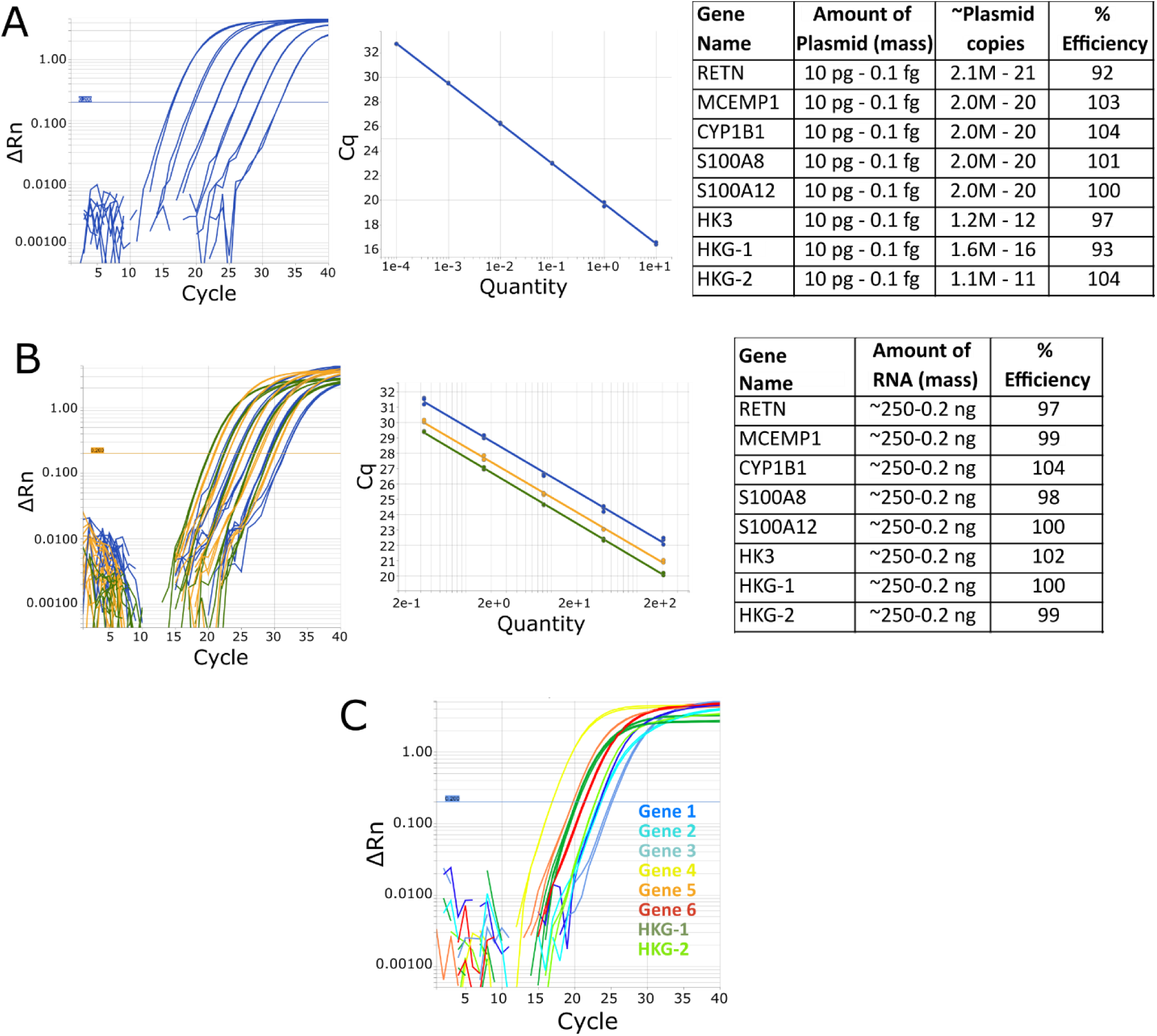
– Development and performance summary of RT-qPCR primers and probes used in the Sepset signature. PCR primers were developed for each sepsis gene signature gene as well as two HKGs and their reaction efficiency was evaluated in a single-plex reaction using gene specific plasmids as the reaction template (A). Shown is the representative amplification curves and corresponding standard curve for the MCEMP1 target along with a table summarizing the reaction efficiencies across all genes. In all cases, the efficiency was close to 100% and consistent RT-qPCR results could be obtained with as low as 10-20 copies of template in the reaction. The PCR primers were then combined in triplex reactions (B) and their reaction efficiencies were determined using RNA isolated from healthy donor blood (n=4 replicates). Shown is a representative amplification curve and corresponding standard curve for one of the multiplex reaction wells. Reaction efficiencies for all the RT-qPCR reactions were close to 100% and consistent results could be obtained with as little as ∼0.2 ng of RNA used as input for the RT-qPCR reaction. A representative amplification curve for the RT-qPCR assay showing the expression pattern of all six signature genes as well as the two housekeeping genes is also shown (C).

**Supplemental Figure S4.**
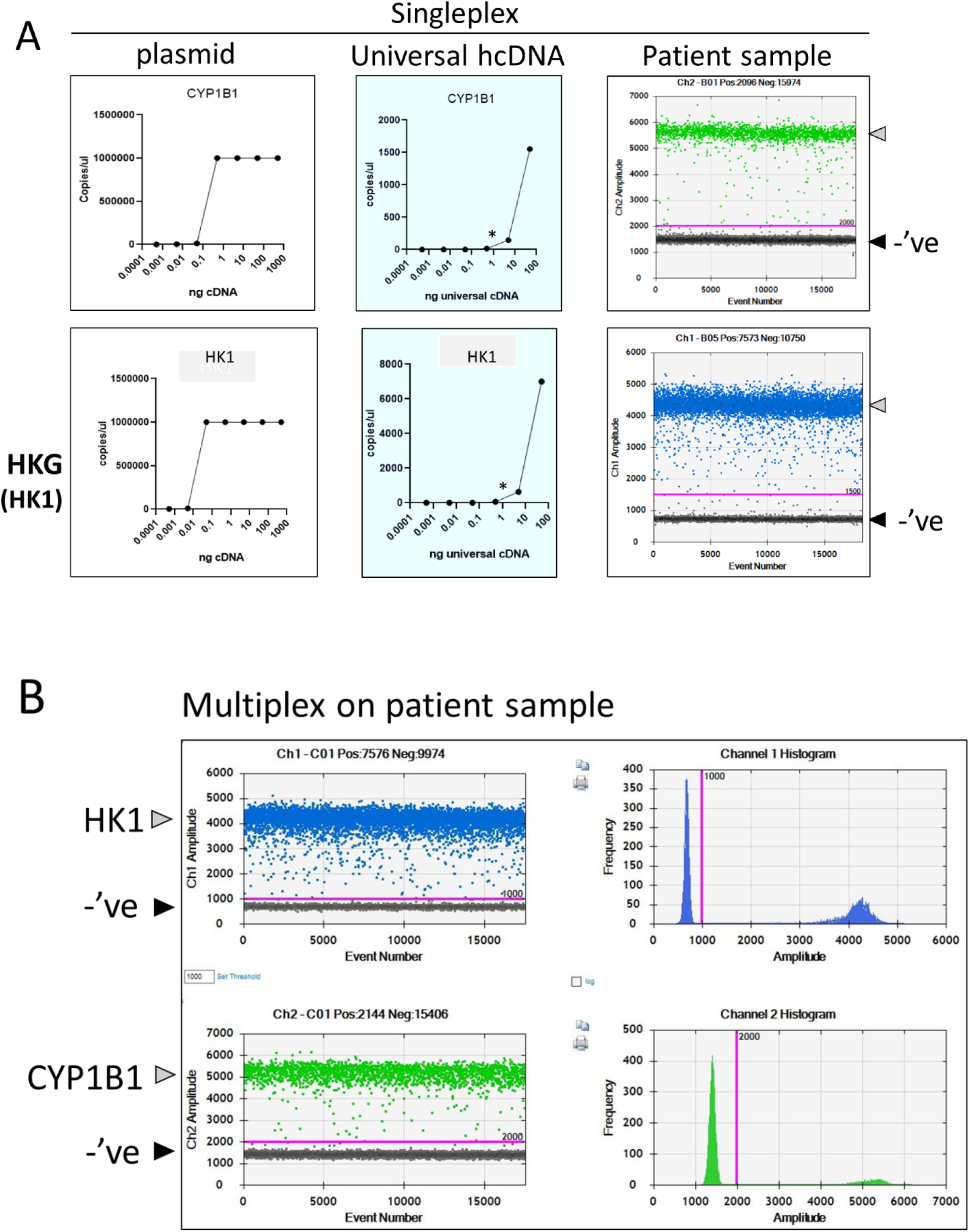
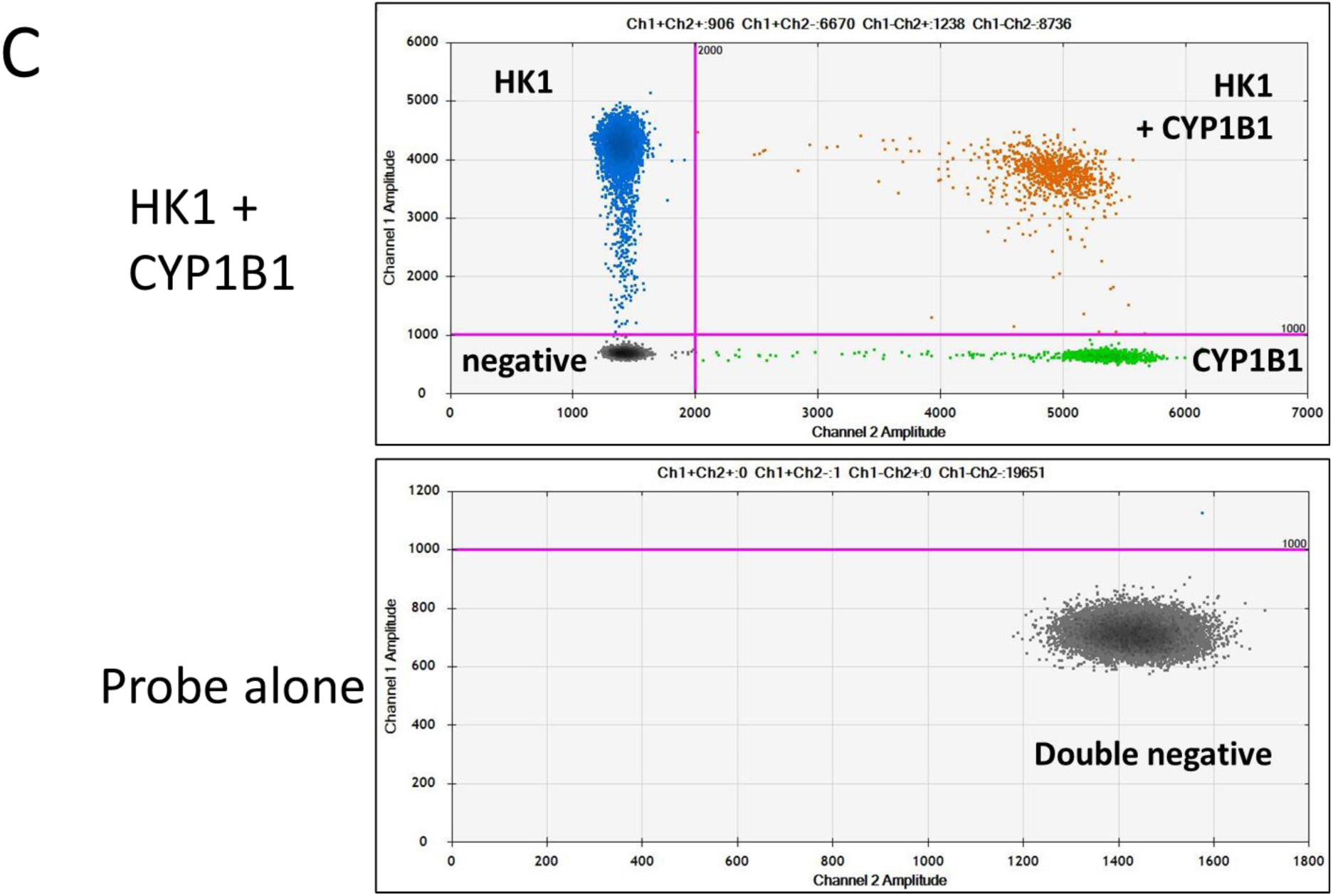
Benchmarking of Sepset signature probes. A. (A. Left panels) Gene specific plasmids were obtained and purified and used as template for the probes using the BioRad AutoDG and QX200 ddPCR system (“gold standard”). Shown are CYP1B1 as a representative for the genes of interest (top) and PTP4A2 as representative housekeeping gene (HKG, bottom). Serial dilutions of the plasmid were used to determine LOI and dynamic range of the probes. (A. Middle panels) Serial dilutions of universal human cDNA were used as template for “gold standard” ddPCR to determine the concentration of input (500 pg) for optimal profiling of patient samples. (A. Right panels) Representative single-plex (CYP1B1 and PTP4A2) sample tracings from a patient sample showing probe signal (positive partition) and negative partition at optimal (500 pg) input concentration. B. Probes were utilized in duplex using (500 pg) patient samples and counts were assessed from each patient, in triplicate. (left panel) Sample tracings showing partitioning of positive and negative droplets. Thresholding was performed on the 2D tracing (B. top right panel) and clear demarcation of double negative droplets (bottom left quadrant), PTP4A2 positive droplets (top left quadrant), CYP1B1 positive droplets (bottom right quadrant) and double positive droplets (top right quadrant) are apparent. Probe alone control rendered only double negative droplets (B. bottom right panel).

**Supplemental Figure S5.**
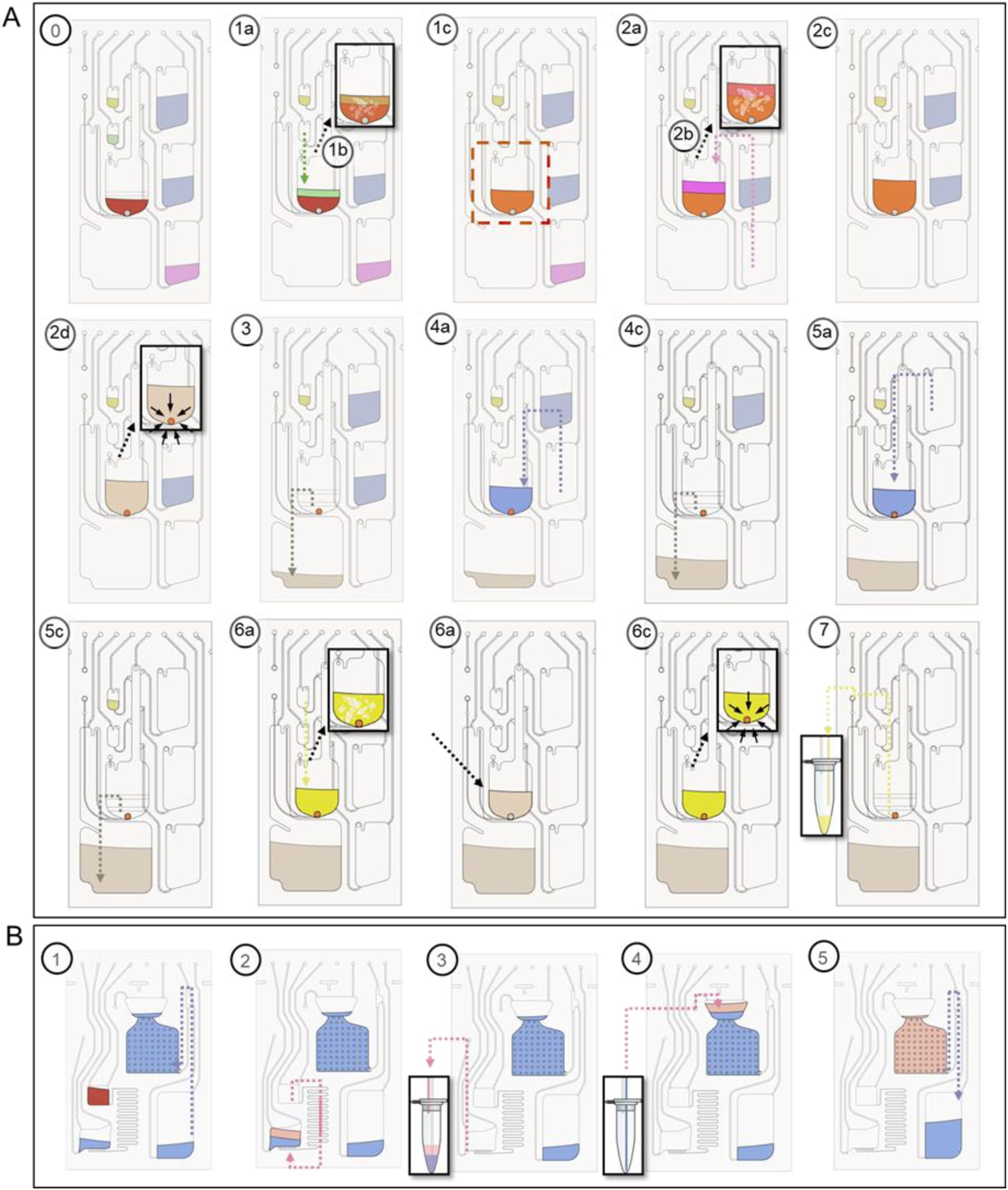
Microfluidic implementation. (A) RNA extraction and (B) ddPCR assays. The platform executes a predetermined sequence of spinning and pneumatic actuation steps to perform the entire analytical protocol in an automated fashion on the cartridge. These steps are represented by the sequence of schematic images showing the status of the cartridge at different steps during the automated RNA extraction protocol and subsequent ddPCR (see text and Table S2 and Table S3 for details). The arrows point in the direction of fluid flow for each step. The activated ports at each step are highlighted by green and red circles, representing the application of positive and negative pressures at that port, respectively.

**Supplemental Figure S6:**
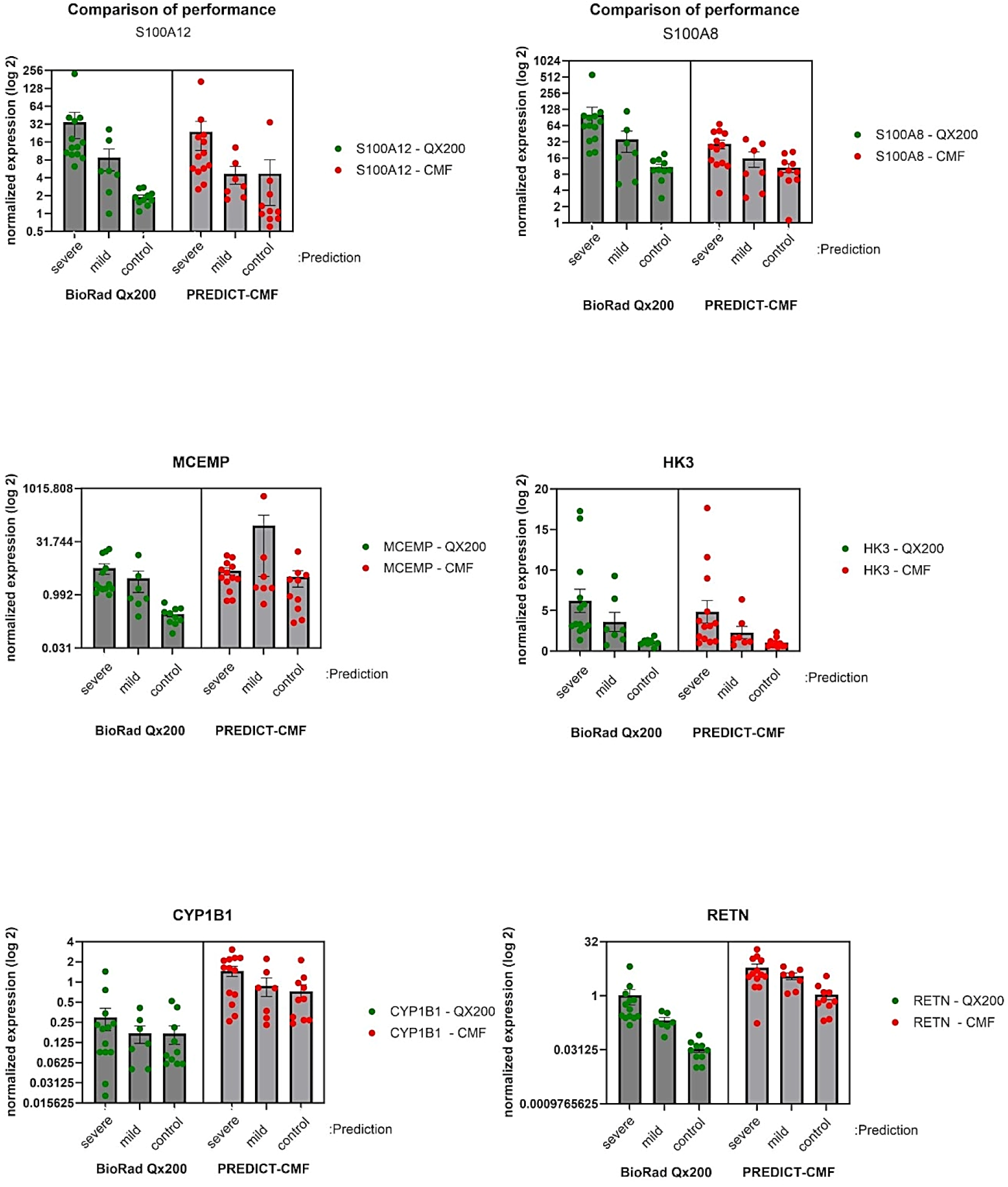
Performance of the PREDICT system compared to BioRad QX200 ddPCR. RNA from the whole blood of ICU patients or healthy controls was isolated, reverse transcribed and the two platforms (BioRad QX200 (green) and PREDICT-CMF (red)) quantitated (copies/µL) the expression of the (6) signature genes of interest (GOI) and (2) housekeeper genes (PTP4A2 and CHTOP). The raw counts of the GOI (labelled at top of plots) were normalized to the GEOMEAN of the HKG [PTP4A2, CHTOP] and were plotted on log_2_ scale. ICU patient samples were classified as “severe or mild” based on their SOFA scores (>2, severe and <2, mild) with the healthy controls.

